# Autoantibodies towards HFE and SYT5 in anti-neutrophil cytoplasm antibody-associated vasculitis relapse

**DOI:** 10.1101/2024.07.25.24310702

**Authors:** Shaghayegh Bayati, Jamsheela Nazeer, James Ng, Michael Hayes, Mark A. Little, Peter Nilsson, Elisa Pin

## Abstract

**Objective:** Identification of those at high and low risk of disease relapse is a major unmet need in the management of patients with ANCA-associated vasculitis (AAV). Precise stratification would allow tailoring of immunosuppressive medication. We profiled the autoantibody repertoire of AAV patients in remission to identify novel autoantibodies associated with relapse risk.

**Methods:** Plasma samples collected from AAV patients in remission were screened for novel autoantibodies using in-house generated protein arrays including 42,000 protein fragments representing 18,000 unique human proteins. Patients were categorized based on the occurrence and frequency of relapses. We modelled the association between these antibodies and relapse occurrence using descriptive and high dimensional regression approaches.

**Results:** We observed nine autoantibodies at higher frequency in samples from AAV patients experiencing multiple relapses compared to patients in long-term remission off therapy (LTROT). LASSO analysis identified six autoantibodies that exhibited an association with relapse occurrence after sample collection. Antibodies targeting HFE and SYT5 were identified as associated with relapse in both analyses.

**Conclusion:** Through a broad protein array-based autoantibody screening, we identified two novel autoantibodies as candidate biomarkers of relapse in AAV.

**Key messages of this study:**

1. Our multi-step screening based on high-throughput and high-multiplexing protein arrays allowed to identify novel autoantibodies in AAV patients.
2. Our study identified two new autoantibodies as candidate biomarkers for predicting AAV patients at risk of relapse.
3. The risk of relapse may be better reflected by the presence of specific autoantibodies than by the overall autoantibody load in patients with AAV.

## 1. Introduction

Anti-neutrophil cytoplasm antibody (ANCA) -associated vasculitides (AAV) is a group of rare systemic autoimmune disorders characterized by inflammation and necrosis of the small blood vessels causing damage potentially to any organ (1). Clinicopathological presentations of AAV include granulomatosis with polyangiitis (GPA), characterized by granulomatous inflammation and often involving the respiratory tract, microscopic polyangiitis (MPA), vasculitis with absence of granulomas and frequent kidney involvement, and eosinophilic granulomatosis with polyangiitis (EGPA), rarer compared to MPA and GPA and involving several organs including lungs and skin (2). Circulating ANCAs are a characteristic trait of AAV and play a key role in AAV pathogenesis by binding to components of the neutrophil and monocyte granules, causing their dysregulated activation. The two main antigens recognized by ANCAs are myeloperoxidase (MPO) and proteinase 3 (PR3). Anti-MPO antibodies are typically associated with MPA, while anti-PR3 antibodies are typically associated with GPA (3, 4).

AAV is characterized by a relapsing-remitting course, with alternation between asymptomatic phases and symptomatic disease flare. Predicting which patients are more at risk of relapse is one of the major unmet needs in AAV clinical management, as it would help to modulate immunosuppression and minimise the risk of over/under-treatment. Besides their use as serological markers for AAV diagnosis, ANCA titres have shown to correlate with disease activity (5-9). C-reactive protein (CRP) and erythrocyte sedimentation rate (ESR) are also used as markers of active AAV in untreated patients at first presentation. However, none of these markers has shown sufficient fidelity in predicting relapses (10).

Autoantibodies have already been reported as good candidate biomarkers for relapse prediction in other autoimmune diseases (11-13). With the aim of finding novel predictive biomarkers in AAV, we designed and performed a broad autoantibody screening in patients at remission with follow-up data using an in-house developed protein array (14-16).

## 2. Materials and Methods

### 2.1 Study group

Inclusion criteria were a definite diagnosis of AAV according to the EMA small vessel vasculitis algorithm (17) and excluding patients with a co-existent anti-glomerular basement membrane antibody enrolled in the Irish National Rare Kidney Disease Registry and Biobank between 2012-2020 (18). Patients’ demographic and information regarding diagnosis (MPA, GPA, EGPA), ANCA serology across the disease, overall organ involvement, vital status at last follow up, and cause of death is provided (**Tab. 1 and Suppl. Tab. 1**). Moreover, information specific for the time of sampling is provided and includes time from diagnosis, time to death (when appropriate), estimated glomerular filtration rate (eGFR), CRP, ANCA status, and treatment received (**Tab. 2**). ANCA serotype was tested in each sample by indirect immunofluorescence (IIF) as well as enzyme-linked immunosorbent assay (ELISA). For each patient, one plasma sample was retrieved during a period of disease remission (defined as Birmingham Vasculitis Activity Score = 0) (19) of at least 6 months duration. The relapse status was established at both patient- and encounter-level. The patient-level and encounter-level classifications were both applied in this study to identify the most promising autoantibody associations with relapse and to mitigate against over-fitting. At encounter-level, post-sample relapse status was determined by whether a relapse (defined as BVAS score >0) occurred after the time of sample collection (“post-sample relapsers”). At the patient-level we focused on two subgroups with extreme phenotypes. The first group included patients in remission for 3 years or longer who were receiving no immunosuppressive medication apart from corticosteroid equivalent to prednisolone 5 milligrams or less, and was therefore termed “long-term remission off therapy” (LTROT). The other group included patients with at least 3 relapse events (frequent relapsers) (**Tab. 3**). This study was approved by the SJH/TUH Research Ethics Committee (2020-05 List 17 – Amendment) and informed consent was collected for all study participants.

### 2.2 Study design

Autoantibody profiling was performed through three-phase screening (**Fig. 1**). In phase 1, we screened IgGs in four plasma pools representing anti-MPO positive LTROT, anti-PR3 positive LTROT, anti-MPO positive frequent relapsers, and anti-PR3 positive frequent relapsers by using in-house generated planar antigen arrays including 42,000 protein fragments representing 18,000 unique human proteins. In phase 2, targets selected by planar array test were merged with a selection of proteins from the literature to generate a bead array that was used to perform a pilot targeted screening of plasma samples from 13 LTROTs and 8 frequent relapsers. Further selected antigens from this screening were used to generate a new bead-array that we applied in phase 3 to test the full sample set including 246 plasma samples.

**Figure 1.**
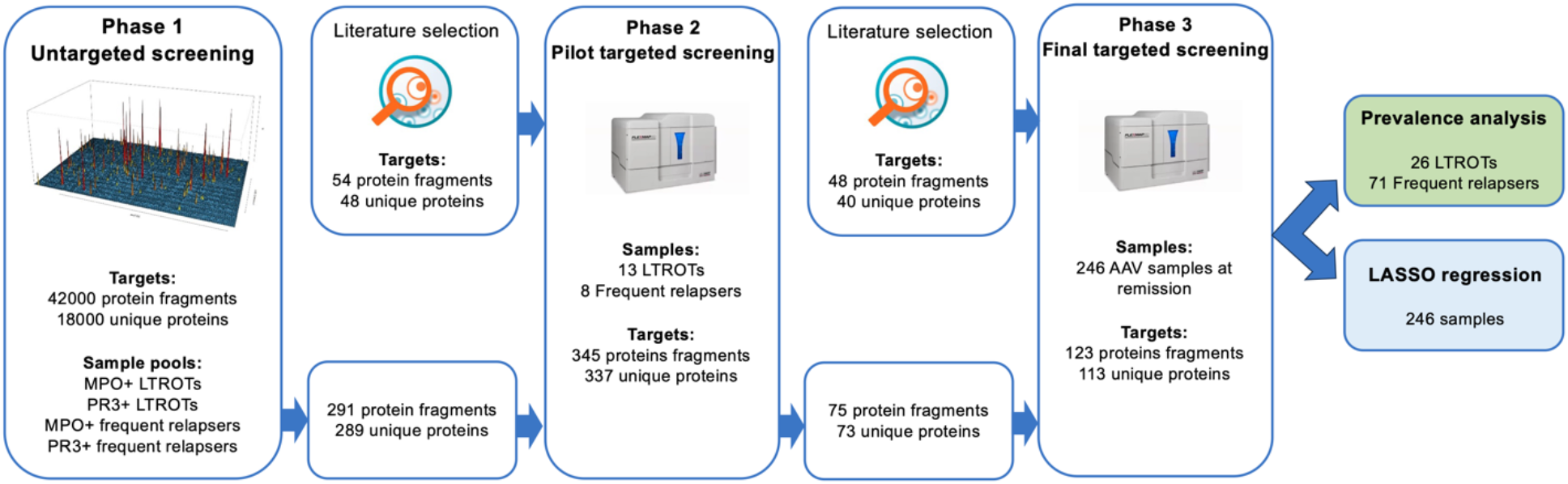
Study design. The figure represents the workflow of the multi-step autoantibody screening.

The data generated from the full sample set targeted screening was submitted to two parallel analyses. We first determined and compared the autoantibody prevalence in LTROTs versus frequent relapsers, and then applied LASSO regression model on the whole sample set (n=246) to identify autoantibodies associated with post-sample relapse occurrence.

### 2.3 Antigens

Protein fragments (40–100 amino acids) produced within the Human Protein Atlas project served as antigens in this study (www.proteinatlas.org). Each protein fragment was designed to represent the part of the gene coding region with the least sequence homology with other human genes. These gene sequences were cloned into expression vectors used for antigens production in E. coli (20).

### 2.4 Untargeted autoantibody screening on planar antigen array – Phase 1

Our in-house generated antigen planar array was applied to test four plasma pools representing patients classified as anti-MPO positive LTROT, anti-PR3 positive LTROT, anti-MPO positive frequent relapsers, and anti-PR3 positive frequent relapsers (**Fig. 1**). Each pool was generated by mixing equal amounts of three plasma samples from three AAV patients. The assay was carried out as previously described (21). Pools were diluted 1:33.3 (final dilution of each single sample is then 1:100) in PBS 0.1% (v/v) Tween20 (Thermo Fisher Scientific, Waltham, MA, USA), 3% bovine plasma albumin (Saveen Werner, Limhamn, Sweden), 5 % (v/v) skim-milk powder (Sigma-Aldrich, St. Louis, MO, USA), and 160 μg/mL His_6_ABP, and incubated for 15 minutes. This allows us to block antibodies reactive to the His_6_ABP tag (i.e., a six-histidine stretch and a streptococcal albumin binding protein) included in each protein fragment for purification purposes. After the pre-incubation, 100 μl of diluted sample was added on the array and incubated for 1 hour. After washing the sample excess, the array was incubated with 0.5 μg/mL hen anti-His6ABP IgY (Immunotech HPA, Stockholm, Sweden). Detection was performed with 130 ng/mL goat anti-chicken IgY Alexa Fluor® 555 (A21437, Invitrogen, Waltham, MA, USA) and 130 ng/mL goat anti-human IgG (H + L) Alexa Fluor® 647 (A21445, Life Technologies, Carlsbad, CA, USA), followed by readout with InnoScan® 1100 laser scanner (Innopsys, Chicago, IL, USA). GenePix Pro 5.1 (Molecular Devices LLC, San Jose, CA, USA) was used for image analysis.

### 2.5 Targeted autoantibody screening using antigen bead array: Phase 2/3

Targeted autoantibody screenings were performed with antigen bead-arrays (**Fig. 1**). In the pilot targeted screening we tested plasma samples from 13 LTROT and 8 frequent relapser AAV patients using a bead-array including 345 protein fragments representing 337 unique proteins (**Suppl. Tab. S2**). In the final targeted screening, we tested the whole sample set of 246 plasma samples using a smaller bead array comprising 123 antigens representing 113 unique proteins (**Suppl. Tab. S3**). The assays were carried out as previously described (14). Briefly, samples were diluted 1:250 in PBS-T 0.05%, 3% (w/v) BSA, 5% (w/v) skim-milk powder, supplemented with 160 μg/ml His6ABP), pre-incubated for 1 hour to block any anti-tag antibodies, then applied on the bead-array for 2 hours. Antigen-autoantibody immunocomplexes were stabilized using with 0.2% paraformaldehyde. Detection was performed with 0.4 μg/mL R-PE conjugated goat anti-human IgG (H10104, Invitrogen, Waltham, MA, USA), followed by read-out using Luminex FLEXMAP 3D® (Luminex Corp., Austin, TX, USA).

### 2.6 Data analysis

#### 2.6.1 Planar and bead-array data normalization and binarization

The R program version 4.3.1 was used for the data analysis. The fluorescence intensity signals (raw data) measured by the InnoScan® 1100 laser scanner (planar array) and the Luminex FLEXMAP 3D® (bead array) were normalized to eliminate the effect of sample specific background noise and transformed into enrichment scores. Specifically, the planar array raw data was normalized and transformed by using the formula: nSD = (xi-mean(xn))/SD(xn), where nSD represented the number of standard deviations above the array-specific mean intensity, xi was the raw intensity signal of the single protein fragment on the array, means(xn) was the mean of the raw intensity signals across the array, and SD(xn) was the standard deviation across the array. After normalization, we set a cutoff of 4SD to select antigens to be passed on to further testing by bead-arrays. The bead-array raw data was normalized and transformed by using the formula nMAD = (xi-median(xn))/MAD (xn), where xi was the raw intensity signal of each single protein fragment in a sample, median(xn) was the median intensity signal across all antigens in a sample, MAD (xn) was the median absolute deviation across all antigens in a sample. A cut-off was defined based on the autoantibodies signal distribution across all samples and data was binarized. A value 1 was assigned to signals passing the cutoff and the sample was therefore deemed *positive* for IgG targeting the specific antigen. Signals below the cut-off were assigned a 0 value. For each sample, the total number of autoantibodies was evaluated by summing the number of signals passing the cut-off.

#### 2.6.2 Comparison of autoantibody prevalence in LTROTs versus frequent relapsers

R version 4.3.1 was used for the analysis. The analysis was performed on data from the pilot screening and full-cohort targeted screenings. Wilcoxon test was applied to compare the total number of autoantibodies in LTROTs and frequent relapsers, with the aim to evaluate whether frequent relapses are associated with a general increase in the number of autoantibodies. We assessed the prevalence of each single autoantibody in LTROTs and frequent relapsers using Fisher’s exact test, with the objective of identifying specific autoantibodies at higher prevalence in frequent relapsers. Heatmaps and cluster analysis were generated to evaluate the distribution of selected autoantibodies across patient groups.

#### 2.6.3 Autoantibody association with future relapse

LASSO regression analysis using R version 4.2.2 was conducted to identify antibodies associated with future relapse. LASSO (Least Absolute Shrinkage and Selection Operator) is a regression analysis method that combines variable selection and regularization (22-24). LASSO adds a penalty term for the absolute values of the regression coefficients, effectively setting the coefficients of less relevant variables to zero. Consequently, LASSO performs automatic variable selection and simplifies the regression model by focusing only on the most important variables.

Different combinations of raw, normalized and binary data from the full cohort targeted screening plus 19 baseline features led to 27 analyses (**Suppl. Tab. S4 and S5**). The outcome variable was included in the model as binary (relapse or no relapse), continuous (number of relapses per year), and time-to-event (time until the first relapse event). For binary outcomes, LASSO logistic regression models were employed. LASSO linear regression models were used for continuous outcomes, while LASSO Cox regression models were applied for time-to-event outcomes. The LASSO penalty could be applied to all features or solely to autoantibody features. In each of the 27 LASSO regressions, we focused on autoantibodies with positive estimated coefficients that were deemed associated with post-sample relapse.

## 3. Results

### 3.1 Description of cohort

The study cohort comprised 246 patients (55% male) with one sample each collected at remission (BVAS=0) **(Tab. 1-3, Suppl. Fig. 1)**. Patients with MPA were older than those with GPA (59 years and 51 years, respectively). Eight patients were ANCA negative, and one patient was positive for both anti-MPO and anti-PR3 antibodies. At the time of the study, follow-up data was available for all patients, with majority of the patients (84%) reported as alive. The most frequently affected organs were kidneys (83%), lungs (53%) and ear-nose-throat (46%).

### 3.2 Phase 1 and 2

Through phase 1 of the study, 291 protein fragments representing 289 unique proteins were selected for further analysis. These and 48 more proteins (54 protein fragments) selected from literature were included in phase 2, where IgG reactivity was detected towards 56% of the tested antigens (192 out of the 345). Each autoantibody was detected in 1 to 7 samples, while in each sample was positive for 3 to 41 autoantibodies. We observed a higher prevalence of autoantibodies binding to activating transcription factor 3 (ATF3), methyltransferase-like protein 6 (METTL6), potassium channel subfamily K member (KCNK4), and BMERB domain containing 1 (BMERB1) (p-value < 0.05) in relapsing patients compared to LTROT (**Suppl. Fig. S2**). These four protein fragments, together with 71 additional fragments (69 unique proteins) that were reactive in more than 3 samples, were moved on for further verification in phase 3.

### 3.3 Phase 3

#### 3.3.1 LTROT versus frequent relapsers

Ninety-seven of the 246 tested samples were classified as either LTROT (n=26) or frequent relapsers (n=71). As in phase 2, similar autoantibody load was detected in females and males and between LTROTs and frequent relapsers. No correlation was observed between autoantibody load and age. On the other hand, anti-MPO positive patients showed a trend towards higher autoantibody load compared to anti-PR3 positive patients, and in MPA patients compared to GPA patients (**Fig. 2**).

**Figure 2.**
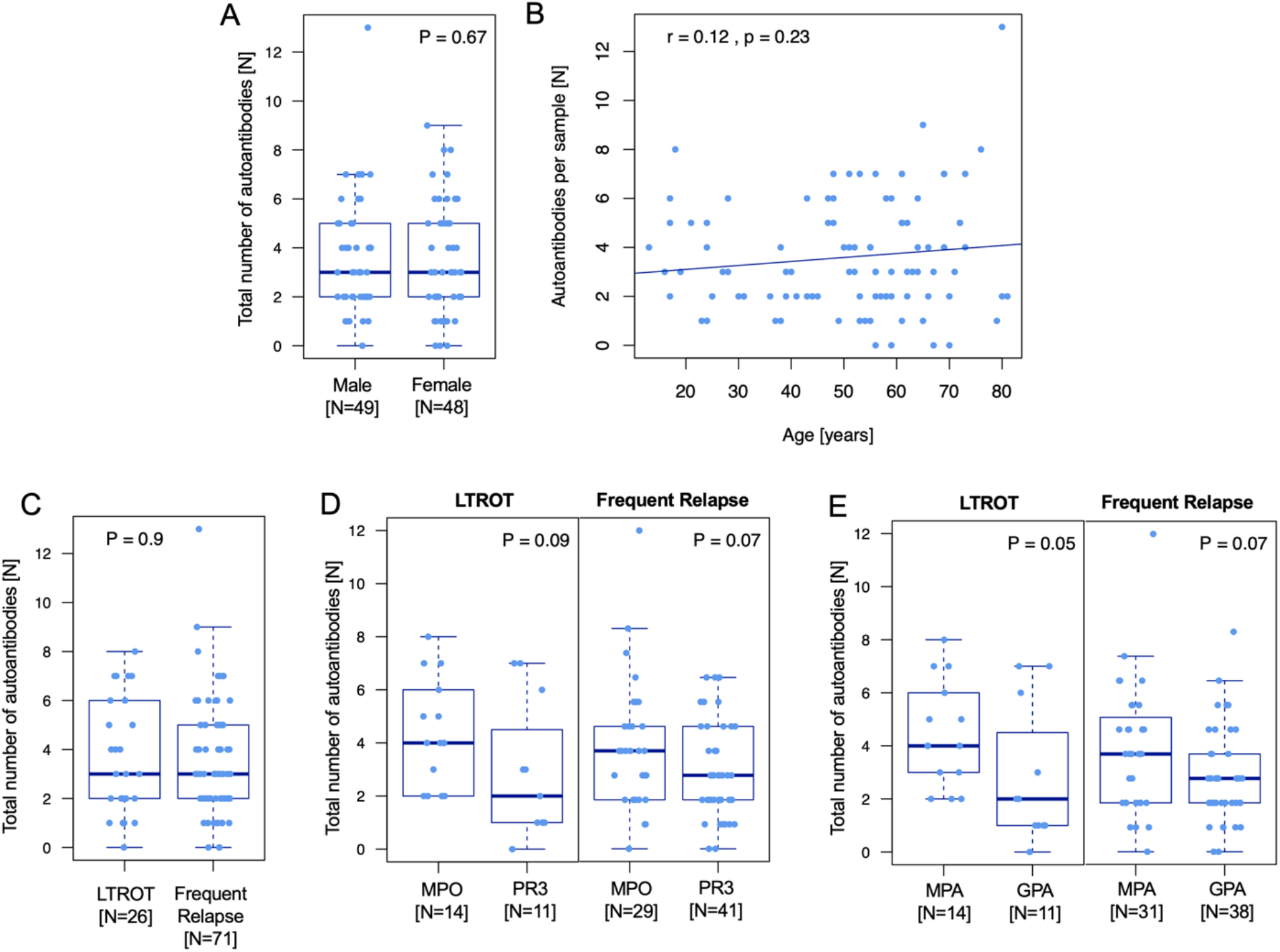
Autoantibody load in LTROTs versus Frequent Relapsers. A) Total number of autoantibodies in female and male patients. B) Correlation between total number of autoantibodies and age. C) Total number of autoantibodies in LTROT and frequent relapsers. D) Total number of autoantibodies in LTROTs and frequent relapsers classified by ANCA serology. Two ANCA negative patients were not included. E) Total number of autoantibodies in LTROTs and frequent relapsers classified by diagnosis. The plot focuses on MPA and GPA, while the three EGPA patients were not included due to low numerosity.

Nine fragment-specific autoantibodies showed high-intensity signals and at least 5 percentage units’ higher prevalence in frequent relapsers **(Fig. 3)**. Among them, anti-ATF3 and anti-METTL6 antibodies were confirmed from the Phase 2. The remaining seven bind to synaptotagmin 5 (SYT5), homeostatic iron regulator (HFE), chromosome 11 open reading frame 54 (C11orf54), transmembrane protein 123 (TMEM123), transforming growth factor beta receptor 3 (TGFBR3), integrin beta 1 (ITGB1) and kinesin family member 15 (KIF15). A total of 40 of the 71 relapsing patients (56%) and 4 of 26 LTROT patients (15%) were positive for at least one of the nine autoantibodies (p-value = 0.0004). Moreover, 21% of frequent relapsers (15/71) and none of the LTROT AAV patients were positive for at least 2 autoantibodies (p = 0.0093). ROC curve analysis resulted in Area Under the Curve (AUC) = 71.4%. When normalized intensity [AU] was used as input, the best test performance for the 9-antigen panel was achieved at 45.5 AU, with 66.2% sensitivity and 80.8% specificity, AUC = 73% **(Suppl. Fig. S3A-D**).

**Figure 3.**
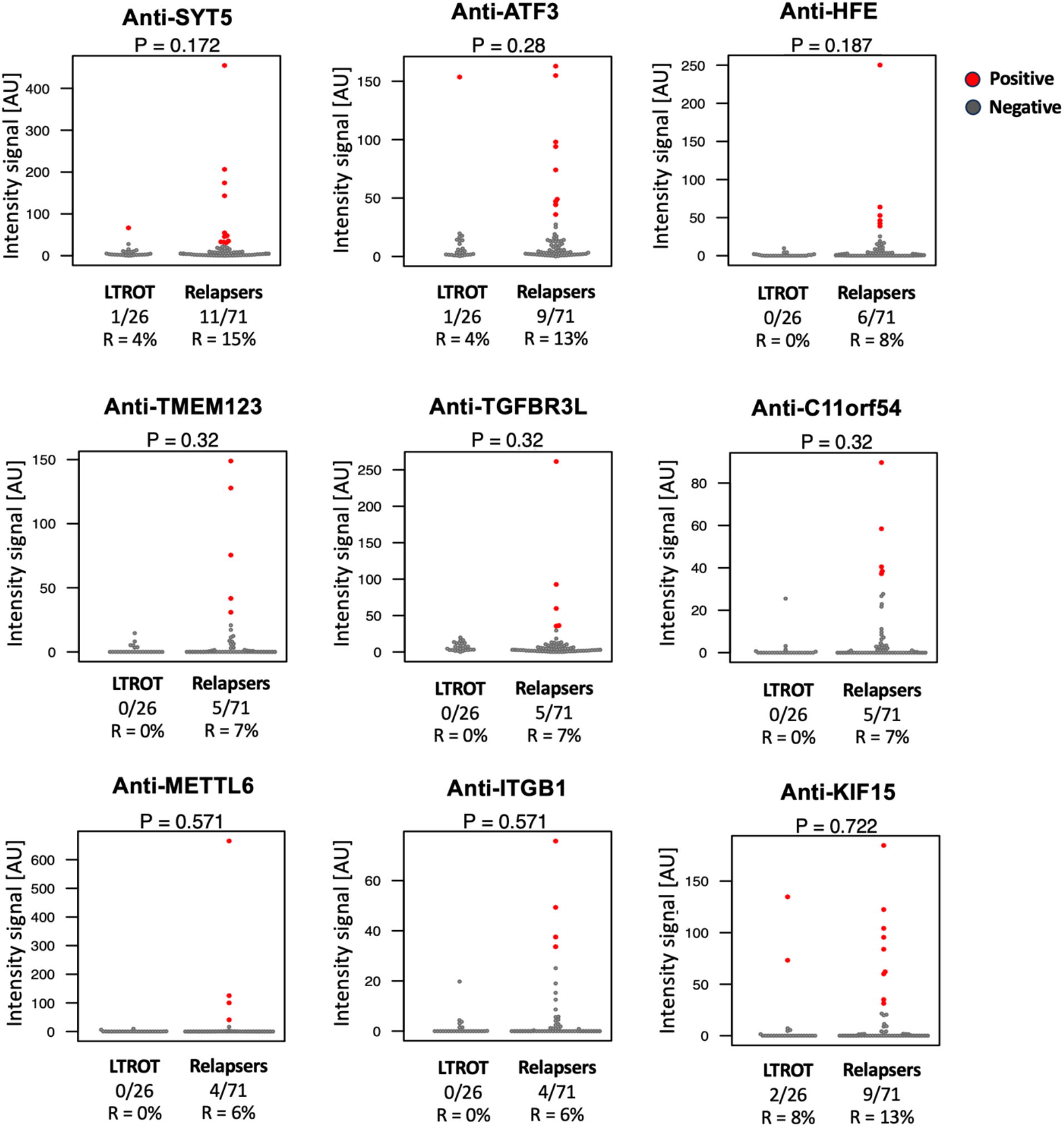
Autoantibodies with higher prevalence in frequent relapsers patients. Each dot in the plot represents a patient. Intensity signals refer to normalized intensity as described in the materials and methods section. Patients seropositive for the specific autoantibody are shown in red. R = % of reactive/seropositive samples. P-values refer to Fisher’s exact test.

#### 3.3.2 Autoantibodies correlating with post-sample relapse

The LASSO regression analysis revealed six autoantibodies with positive coefficients in at least one of the 27 LASSO analyses (**Fig. 4A**). These six autoantibodies, targeting T cell leukemia homebox 2 (TLX2), Golgi associated RAB2B interactor family member 3 (FAM71B), FXYD domain containing ion transport regulator 2 (FXYD2), homeostatic iron regulator (HFE), Ras-related protein Rab-7a (RAB7A), and synaptotagmin 5 (SYT5) were associated with post-sample relapse (**Fig. 4B)**. Anti-TLX2 displayed positive coefficient values in 8 regression analyses, making it the autoantibody most strongly associated with post-sample relapse. Notably, anti-SYT5 and anti-HFE were also included among the nine autoantibodies selected in the Phase 3 LTROT versus frequent relapser analysis. We therefore consider these overlapping results as the most promising autoantibody associations with relapse.

**Figure 4.**
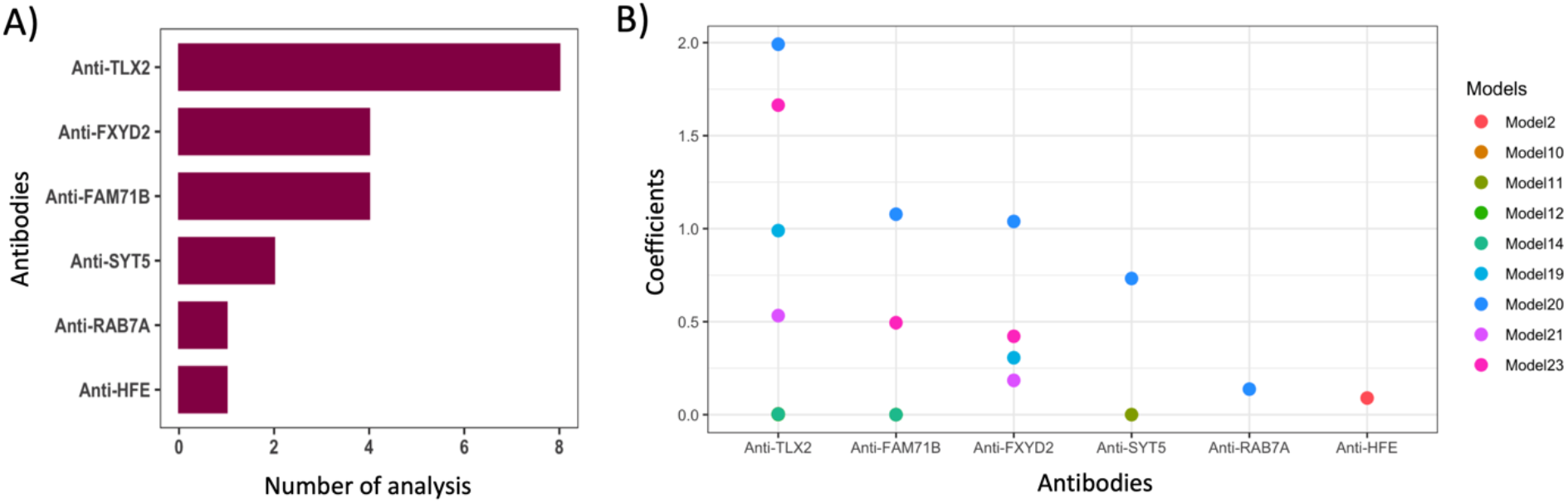
LASSO regression analysis. The analysis included the full cohort of 246 samples and 27 different models listed in supplementary table S3. (A) List of 6 significant antibodies and the number of analyses in which they have positive coefficient values. (B) Presentation of the 6 antibodies with positive coefficient values across various analyses. 27 different models are used in the study. The details are provided in the Supplementary table S4 and 5.

## 4. Discussion

The aim of this study was to identify novel autoantibodies associated with relapse in AAV patients. Predicting relapses in these patients is important in the attempt to minimize organ damage and reduce the risks related to adverse events and infections caused by immunosuppressive treatment (25). Autoantibodies are a hallmark of autoimmune diseases and have demonstrated their usefulness as diagnostic and prognostic biomarkers (26), and therefore represent good candidates for relapse prediction. We performed a broad IgG screen of plasma samples collected from 246 AAV patients at remission by using protein arrays generated with protein fragments from the Human Protein Atlas collection (27, 28). The main results showed nine autoantibodies present in subgroups of patients at high risk of relapse (frequent relapsers) and six autoantibodies associated with future relapse. Two autoantibodies were identified by both analyses and were therefore considered as the most promising relapse associated autoantibodies: these autoantibodies target the homeostatic iron regulator (HFE) and synaptotagmin 5 (SYT5) proteins. These are integral membrane proteins characterized by a single transmembrane alpha-helix and exposed on the extracellular (HFE) and extravesicular (SYT5) compartments (**Suppl. Fig. 4**). The protein fragments used in our study represent parts of the extracellular or extravesicular protein domains, therefore including particularly accessible epitopes. To the best of our knowledge, neither of these proteins have been previously reported in connection to AAV or linked to relapse in AAV or other autoimmune diseases.

SYT5 is a protein involved in calcium-dependent exocytosis (29) and enriched in the brain, where it is mainly involved in neuronal signaling, but also expressed in endothelial cells (30). Exocytosis represents the initial response necessary to restore the endothelial barrier after injury, by initiating platelet rolling and thrombosis (31, 32). Endothelial injury is one of the hallmarks of AAV, and it is mainly induced by neutrophils interacting with the endothelium (33). Autoantibodies binding to SYT5 may therefore capture the attempted repairing efforts of endothelial cells which is ongoing in frequent relapsers, or which precedes a new relapse event.

The HFE protein is expressed in most human tissues and cell types, including immune cells and especially monocytes and neutrophils (34). HFE shows homology with major histocompatibility complex (MHC) class I and is potentially involved in cross-talk with the MHC Class 1 antigen presentation pathway (35). The protein plays an important role in the iron metabolism (36). Iron deficiency is a common cause of anemia, a frequent complication in renal disease and AAV (37, 38). While the significance of autoantibodies targeting HFE and their link to relapse needs to be clarified, the involvement of HFE in the iron metabolism and its possible cross-talk with an antigen presentation pathway makes of it an interesting target to further explore.

To confirm the association with relapse and prediction potential of these two antibodies, independent and larger sample cohorts need to be analyzed. Further functional studies are also needed to clarify whether the presence of these autoantibodies is to be considered as an epiphenomenon or whether they play a pathological role.

Our results do not show any increase in the autoantibody load for patients at high risk of relapse (frequent relapsers) compared to LTROT patients, suggesting that the composition of the autoantibody profile may be more informative in terms of relapse prediction potential. On the other hand, MPA patients showed a trend with higher number of autoantibodies compared to GPA and, as expected, a similar trend was identified for anti-MPO compared to anti-PR3 patients. This trend was recently reported in another study on AAV (39) and could be possibly explained by differences on an immunoreactive level between the different clinicopathological subgroups and ANCA serotypes of AAV (40, 41).

This study presents limitations. First, we analyzed plasma samples collected at a single remission time point. Future validation studies should involve independently collected longitudinal cohorts including samples collected at diagnosis (possibly pre-treatment), remission and relapse. Ideally, samples representing different remission/relapse cycles should also be included for patients with multiple relapses for a better evaluation of the signal fluctuation for the selected antigens. Dedicated studies are also needed to clarify the impact of different regimens of treatment on autoantibody levels. Moreover, inclusion of samples from the Rare Kidney Disease Biobank caused an unbalanced recruitment with most of the AAV patients presenting with renal involvement. Second, the antigens employed in our study are protein fragments which, by definition, represent only a portion of the protein sequence. This causes loss of information related to epitopes that may be present in the part of the protein sequence that is not covered by the fragment. Moreover, the fragment folding might not necessarily reflect that of the native protein. While this would not represent an issue in the detection of linear epitopes, it may on the other hand limit the representativeness of conformational epitopes. To overcome this limitation and increase the antigen representativeness, future validation studies will include several protein fragments as well as full-length proteins representing the selected targets. Future validation studies should also incorporate MPO and PR3 antigens to enable simultaneous measurement of reactivity towards well-established and novel antigens using the same assay platform. Unfortunately, this was not feasible in the presented study because the available MPO and PR3 protein fragments do not encompass the common reactive epitopes identified in AAV (42-44), since these are outside the most unique segment of the protein sequence which was the target of the design of the Human Protein Atlas antigens. To overcome this limitation, MPO and PR3 full-length proteins are currently under evaluation and will be included as positive controls in future assays.

In conclusion, this study identified two autoantibodies possibly linked to risk of relapse in AAV. To the best of our knowledge, these autoantibodies have not previously been identified in relation to AAV and relapse prediction. Verification in independent, longitudinal, and larger cohorts is needed to confirm these associations. Functional studies are also needed to clarify the nature of these autoantibodies and whether they are epiphenomena or may have pathogenic role. Once validated in independently collected sample cohorts, the array-based assay could be translated into ELISA tests to allow for an easier and possible future translation of the test into the clinical practice (39).

## Data Availability

Subject level Data and codes underlying this article cannot be shared publicly as it contains sensitive personal information which is protected by the GDPR. The data and codes underlying this article will be shared on reasonable request to the SciLifeLab Data Repository for researchers who meet the criteria for access to sensitive personal data.

## Author Contributions

SB: Conceptualization, Investigation, Data curation, Formal analysis, Visualization, Writing – Original draft preparation; JNa: data curation, formal analysis, visualization, Writing – Review and Editing; JNg: data curation, formal analysis, Writing – Review and Editing; ML: Funding acquisition, Conceptualization, Resources, Writing – Review and Editing; MH: Writing – Review and Editing; PN: Funding acquisition, Resources, Supervision, Writing – Review and Editing; EP: Conceptualization, Formal analysis, Data curation, Visualization, Supervision, Writing – Review and Editing.

## Acknowledgements

This study was supported by the European Reference Network for rare immune disorders (RITA).

## Members of the RKD Consortium

Tallaght Hospital: Mark Little*, Peter Lavin**, Catherine Wall**, George Mellotte**, Ronan Mullan**, Jennifer Scott**, Eithne Nic an Ríogh, Ted Fitzgerald**, Hannah O’Keefe**, Rachel Dilworth**, Claire Kennedy**, Limy Wong**, Pamela O’Neill#

St James’ Hospital: Niall Conlon*, Brenda Griffin*, Donal Sexton*

Mater University Hospital: Yvonne O’Meara*, Eoghan White**, Stephen Mahony**

St Vincent’s University Hospital: Eamonn Molloy*, John Holian**, Michael Hayes**

Galway University Hospital: Matthew Griffin*, David Lappin**, Conor Judge**, Sarah Cormican**, Blathnaid O’Connell**, Michelle Clince**

Limerick University Hospital: Liam Casserly*, Edward McGonagle**

Cork University Hospital: Sarah Moran*, Michael Clarkson*, Alyssa Verrelli**, Sinead Stoneman**, Fergus Daly**, Laura Slattery**, Aisling Murphy#

Beaumont Hospital: Mark Little*, Declan De Freitas**, Peter Conlon**, Mark Denton**, Carol Treanor**, Colm Magee**, Conall O Seaghdha**, Ciara Magee**, Paul O’Hara**, Susan McGrath**, Brona Moloney**, Dean Moore**, Dearbhla Kelly**, Mary McCarthy**, Tamara Wanigasekera**, Ayanfeoluwa Obilana** Dervla Connaughton**

*Principal Investigator; **Co-investigator; #Study Coordinator

## Ethics

The study was conducted in accordance with the Declaration of Helsinki and approved by the SJH/TUH Research Ethics Committee (ref. nr. 2020-05 List 17 – Amendment). Informed consent has been obtained from the subjects participating in this study.

## References

1. Almaani S, Fussner LA, Brodsky S, Meara AS, Jayne D. ANCA-Associated Vasculitis: An Update. J Clin Med. 2021;10(7).

2. McKinney EF, Willcocks LC, Broecker V, Smith KG. The immunopathology of ANCA-associated vasculitis. Semin Immunopathol. 2014;36(4):461–78.

3. Robson JC, Grayson PC, Ponte C, Suppiah R, Craven A, Judge A, et al. 2022 American College of Rheumatology/European Alliance of Associations for Rheumatology Classification Criteria for Granulomatosis With Polyangiitis. Arthritis Rheumatol. 2022;74(3):393–9.

4. Pyo JY, Ahn SS, Song JJ, Park YB, Lee SW. Reclassification of previously diagnosed GPA patients using the 2022 ACR/EULAR classification criteria. Rheumatology (Oxford). 2023;62(3):1179–86.

5. Hagen EC, Daha MR, Hermans J, Andrassy K, Csernok E, Gaskin G, et al. Diagnostic value of standardized assays for anti-neutrophil cytoplasmic antibodies in idiopathic systemic vasculitis. EC/BCR Project for ANCA Assay Standardization. Kidney Int. 1998;53(3):743–53.

6. Cohen Tervaert JW, Damoiseaux J. Antineutrophil cytoplasmic autoantibodies: how are they detected and what is their use for diagnosis, classification and follow-up? Clin Rev Allergy Immunol. 2012;43(3):211–9.

7. Aoyama Y, Inaba T, Takahashi S, Yasuhara H, Hiraoka S, Morimoto T, et al. Antiproteinase 3 antineutrophil cytoplasmic antibody reflects disease activity and predicts the response to steroid therapy in ulcerative colitis. BMC Gastroenterol. 2021;21(1):325.

8. Wallace ZS, Fu X, Liao K, Kallenberg CGM, Langford CA, Merkel PA, et al. Disease Activity, Antineutrophil Cytoplasmic Antibody Type, and Lipid Levels in Antineutrophil Cytoplasmic Antibody-Associated Vasculitis. Arthritis Rheumatol. 2019;71(11):1879–87.

9. Sharma RK, Lovstrom B, Gunnarsson I, Malmstrom V. Proteinase 3 Autoreactivity in Anti-Neutrophil Cytoplasmic Antibody-associated vasculitis-Immunological versus clinical features. Scand J Immunol. 2020;92(5):e12958.

10. Sproson EL, Jones NS, Al-Deiri M, Lanyon P. Lessons learnt in the management of Wegener’s Granulomatosis: long-term follow-up of 60 patients. Rhinology. 2007;45(1):63–7.

11. Delville M, Sigdel TK, Wei C, Li J, Hsieh SC, Fornoni A, et al. A circulating antibody panel for pretransplant prediction of FSGS recurrence after kidney transplantation. Sci Transl Med. 2014;6(256):256ra136.

12. Tun NN, Beckett G, Zammitt NN, Strachan MW, Seckl JR, Gibb FW. Thyrotropin Receptor Antibody Levels at Diagnosis and After Thionamide Course Predict Graves’ Disease Relapse. Thyroid. 2016;26(8):1004–9.

13. Johannet P, Liu W, Fenyo D, Wind-Rotolo M, Krogsgaard M, Mehnert JM, et al. Baseline Serum Autoantibody Signatures Predict Recurrence and Toxicity in Melanoma Patients Receiving Adjuvant Immune Checkpoint Blockade. Clin Cancer Res. 2022;28(18):4121–30.

14. Geroldinger-Simic M, Bayati S, Pohjanen E, Sepp N, Nilsson P, Pin E. Autoantibodies against PIP4K2B and AKT3 Are Associated with Skin and Lung Fibrosis in Patients with Systemic Sclerosis. Int J Mol Sci. 2023;24(6).

15. Haggmark A, Hamsten C, Wiklundh E, Lindskog C, Mattsson C, Andersson E, et al. Proteomic profiling reveals autoimmune targets in sarcoidosis. Am J Respir Crit Care Med. 2015;191(5):574–83.

16. Lourido L, Ruiz-Romero C, Picchi F, Diz-Rosales N, Vilaboa-Galan S, Fernandez-Lopez C, et al. Association of serum anti-centromere protein F antibodies with clinical response to infliximab in patients with rheumatoid arthritis: A prospective study. Semin Arthritis Rheum. 2020;50(5):1101–8.

17. Watts R, Lane S, Hanslik T, Hauser T, Hellmich B, Koldingsnes W, et al. Development and validation of a consensus methodology for the classification of the ANCA-associated vasculitides and polyarteritis nodosa for epidemiological studies. Ann Rheum Dis. 2007;66(2):222–7.

18. Scott J, Nic An Riogh E, Al Nokhatha S, Cowhig C, Verrelli A, Fitzgerald T, et al. ANCA-associated vasculitis in Ireland: a multi-centre national cohort study. HRB Open Res. 2022;5:80.

19. Mukhtyar C, Lee R, Brown D, Carruthers D, Dasgupta B, Dubey S, et al. Modification and validation of the Birmingham Vasculitis Activity Score (version 3). Ann Rheum Dis. 2009;68(12):1827–32.

20. Hober S, Uhlen M. Human protein atlas and the use of microarray technologies. Curr Opin Biotechnol. 2008;19(1):30–5.

21. Jernbom Falk A, Galletly C, Just D, Toben C, Baune BT, Clark SR, et al. Autoantibody profiles associated with clinical features in psychotic disorders. Transl Psychiatry. 2021;11(1):474.

22. Pampel FC, Core SRM. Logistic regression : a primer. 2nd ed. Los Angeles, CA: SAGE Publications, Inc; 2021.

23. Harrell FE, SpringerLink. Regression Modeling Strategies : With Applications to Linear Models, Logistic Regression, and Survival Analysis. 1st 2001. ed. New York ; London New York, NY: Springer Springer New York : Imprint: Springer; 2001.

24. Moore DF, SpringerLink. Applied Survival Analysis Using R. 1st 2016. ed. Cham: Springer International Publishing Springer International Publishing : Imprint: Springer; 2016.

25. Salama AD. Relapse in Anti-Neutrophil Cytoplasm Antibody (ANCA)-Associated Vasculitis. Kidney Int Rep. 2020;5(1):7–12.

26. Ma H, Murphy C, Loscher CE, O’Kennedy R. Autoantibodies - enemies, and/or potential allies? Front Immunol. 2022;13:953726.

27. Uhlen M, Fagerberg L, Hallstrom BM, Lindskog C, Oksvold P, Mardinoglu A, et al. Proteomics. Tissue-based map of the human proteome. Science. 2015;347(6220):1260419.

28. The Human Protein Atlas, v.22.0 2023 [updated 2022-12-07. v. 22.0:[Available from: https://www.proteinatlas.org/.

29. Lenzi C, Stevens J, Osborn D, Hannah MJ, Bierings R, Carter T. Synaptotagmin 5 regulates Ca(2+)-dependent Weibel-Palade body exocytosis in human endothelial cells. J Cell Sci. 2019;132(5).

30. The Human Protein Atlas - SYT5 [Available from: https://www.proteinatlas.org/ENSG00000129990-SYT5.

31. Yamakuchi M, Ferlito M, Morrell CN, Matsushita K, Fletcher CA, Cao W, et al. Exocytosis of endothelial cells is regulated by N-ethylmaleimide-sensitive factor. Methods Mol Biol. 2008;440:203–15.

32. Stolwijk JA, Zhang X, Gueguinou M, Zhang W, Matrougui K, Renken C, et al. Calcium Signaling Is Dispensable for Receptor Regulation of Endothelial Barrier Function. J Biol Chem. 2016;291(44):22894–912.

33. Halbwachs L, Lesavre P. Endothelium-neutrophil interactions in ANCA-associated diseases. J Am Soc Nephrol. 2012;23(9):1449–61.

34. The Human Protein Atlas - HFE [Available from: https://www.proteinatlas.org/ENSG00000010704-HFE.

35. Reuben A, Chung JW, Lapointe R, Santos MM. The hemochromatosis protein HFE 20 years later: An emerging role in antigen presentation and in the immune system. Immun Inflamm Dis. 2017;5(3):218–32.

36. Barton JC, Edwards CQ, Acton RT. HFE gene: Structure, function, mutations, and associated iron abnormalities. Gene. 2015;574(2):179–92.

37. Chang R, Chu KA, Lin MC, Chu YH, Hung YM, Wei JC. Newly diagnosed iron deficiency anemia and subsequent autoimmune disease: a matched cohort study in Taiwan. Curr Med Res Opin. 2020;36(6):985–92.

38. Kawamura T, Usui J, Kaneko S, Tsunoda R, Imai E, Kai H, et al. Anaemia is an essential complication of ANCA-associated renal vasculitis: a single center cohort study. BMC Nephrol. 2017;18(1):337.

39. Mescia F, Bayati S, Brouwer E, Heeringa P, Toonen EJM, Beenes M, et al. Autoantibody Profiling and Anti-Kinesin Reactivity in ANCA-Associated Vasculitis. Int J Mol Sci. 2023;24(20).

40. Samman KN, Ross C, Pagnoux C, Makhzoum JP. Update in the Management of ANCA-Associated Vasculitis: Recent Developments and Future Perspectives. Int J Rheumatol. 2021;2021:5534851.

41. Hilhorst M, van Paassen P, Tervaert JW, Limburg Renal R. Proteinase 3-ANCA Vasculitis versus Myeloperoxidase-ANCA Vasculitis. J Am Soc Nephrol. 2015;26(10):2314–27.

42. Bruner BF, Vista ES, Wynn DM, James JA. Epitope specificity of myeloperoxidase antibodies: identification of candidate human immunodominant epitopes. Clin Exp Immunol. 2011;164(3):330–6.

43. Erdbrugger U, Hellmark T, Bunch DO, Alcorta DA, Jennette JC, Falk RJ, et al. Mapping of myeloperoxidase epitopes recognized by MPO-ANCA using human-mouse MPO chimers. Kidney Int. 2006;69(10):1799–805.

44. Bini P, Gabay JE, Teitel A, Melchior M, Zhou JL, Elkon KB. Antineutrophil cytoplasmic autoantibodies in Wegener’s granulomatosis recognize conformational epitope(s) on proteinase 3. J Immunol. 1992;149(4):1409–15.

