## Supplementary material for "Autoantibodies towards HFE and SYT5 in anti-neutrophil cytoplasm antibody-associated vasculitis relapse"

**Supplementary Table S1. Patients' cause of death.**

| | **Overall n=40** | **GPA n=16** | **MPA n=24** |
|---|---|---|---|
| Cardiac disease, n (%) | 5 (13%) | 2 (13%) | 3 (13%) |
| End stage Renal Disease, n (%) | 1 (2.5%) | 0 (0%) | 1 (4.2%) |
| Infection/Sepsis, n (%) | 13 (33%) | 4 (25%) | 9 (38%) |
| Neurological Disease, n (%) | 3 (7.5%) | 3 (19%) | 0 (0%) |
| Respiratory Disease, n (%) | 1 (2.5%) | 0 (0%) | 1 (4.2%) |
| Cancer, n (%) | 6 (15%) | 4 (25%) | 2 (8.3%) |
| Unknown, n (%) | 11 (28%) | 3 (19%) | 8 (33%) |

**Supplementary Table S2. Pilot targeted screening. Antigen information.**

| **HPRR** | **Gene** | **Gene description** | **Uniprot ID** | **Inclusion criteria** |
|---|---|---|---|---|
| HPRR140351 | EDDM3B | epididymal protein 3B | P56851 | Literature |
| HPRR140757 | MTHFD1 | methylenetetrahydrofolate dehydrogenase, cyclohydrolase and formyltetrahydrofolate synthetase 1 | P11586 | Planar array |
| HPRR140758 | MTHFD1L | methylenetetrahydrofolate dehydrogenase (NADP+ dependent) 1 like;methylenetetrahydrofolate dehydrogenase, cyclohydrolase and formyltetrahydrofolate synthetase 1 | Q6UB35 | Planar array |
| HPRR221275 | SCUBE1 | signal peptide, CUB domain and EGF like domain containing 1 | Q8IWY4 | Literature |
| HPRR260163 | PRICKLE1 | prickle Planar array cell polarity protein 1 | Q96MT3 | Planar array |
| HPRR270049 | ERCC1 | ERCC excision repair 1, endonuclease non-catalytic subunit | P07992 | Planar array |
| HPRR280090 | C3 | complement C3 | P01024 | Literature |
| HPRR280135 | ADA | adenosine deaminase | P00813 | Literature |
| HPRR280163 | C1QA | complement C1q A chain | P02745 | Literature |
| HPRR290194 | FES | FES proto-oncogene, tyrosine kinase | P07332 | Planar array |
| HPRR330007 | IL6 | interleukin 6 | P05231 | Literature |

| | | | | |
|---|---|---|---|---|
| HPRR330247 | NFKB1 | nuclear factor kappa B subunit 1 | P19838 | Planar array |
| HPRR350013 | IL12A | interleukin 12A | P29459 | Literature |
| HPRR360180 | ELK4 | ETS transcription factor ELK4 | P28324 | Planar array |
| HPRR370285 | LMO7 | LIM domain 7 | Q8WWI1 | Planar array |
| HPRR400117 | TSC1 | TSC complex subunit 1 | Q92574 | Planar array |
| HPRR520073 | ZFHX3 | zinc finger homeobox 3 | Q15911 | Planar array |
| HPRR620003 | LTBP1 | latent transforming growth factor beta binding protein 1 | Q14766 | Literature |
| HPRR620023 | IL18 | interleukin 18 | Q14116 | Literature |
| HPRR640050 | NFIB | nuclear factor I B | O00712 | Literature |
| HPRR670216 | TRPM2 | transient receptor potential cation channel subfamily M member 2 | O94759 | Planar array |
| HPRR680141 | CHD4 | chromodomain helicase DNA binding protein 4 | Q14839 | Literature |
| HPRR1440011 | CLPP | caseinolytic mitochondrial matrix peptidase proteolytic subunit | Q16740 | Planar array |
| HPRR1450470 | HSBP1 | heat shock factor binding protein 1 | O75506 | Planar array |
| HPRR1450742 | RBPJ | recombination signal binding protein for immunoglobulin kappa J region | Q06330 | Literature |
| HPRR1820052 | MINDY2 | MINDY lysine 48 deubiquitinase 2 | Q8NBR6 | Planar array |
| HPRR1840161 | MLKL | mixed lineage kinase domain like pseudokinase | Q8NB16 | Planar array |
| HPRR1840197 | EPHB3 | EPH receptor B3 | P54753 | Planar array |
| HPRR1850063 | ALDH1A2 | aldehyde dehydrogenase 1 family member A2 | O94788 | Literature |
| HPRR1920067 | NAPG | NSF attachment protein gamma | Q99747 | Planar array |
| HPRR1950036 | HFE | homeostatic iron regulator | Q30201 | Planar array |
| HPRR1950351 | PON3 | paraoxonase 3 | Q15166 | Planar array |
| HPRR1950646 | LRP11 | LDL receptor related protein 11 | Q86VZ4 | Planar array |
| HPRR1950820 | TNFRSF19 | TNF receptor superfamily member 19 | Q9NS68 | Planar array |
| HPRR1950853 | KLB | klotho beta | Q86Z14 | Planar array |
| HPRR2090170 | CHRM1 | cholinergic receptor muscarinic 1 | P11229 | Planar array |

| | | | | |
|---|---|---|---|---|
| HPRR2150261 | CLDN18 | claudin 18 | P56856 | Planar array |
| HPRR2260138 | UBQLN3 | ubiquilin 3 | Q9H347 | Planar array |
| HPRR2310082 | GDF3 | growth differentiation factor 3 | Q9NR23 | Planar array |
| HPRR2320171 | BNIPL | BCL2 interacting protein like | Q7Z465 | Planar array |
| HPRR2460064 | PDE2A | phosphodiesterase 2A | O00408 | Planar array |
| HPRR2470230 | CAMSAP1 | calmodulin regulated spectrin associated protein 1 | Q5T5Y3 | Planar array |
| HPRR2470419 | WDR31 | WD repeat domain 31 | Q8NA23 | Planar array |
| HPRR2470521 | FREM1 | FRAS1 related extracellular matrix 1 | Q5H8C1 | Planar array |
| HPRR2490002 | ADAMTS2 | ADAM metallopeptidase with thrombospondin type 1 motif 2 | O95450 | Planar array |
| HPRR2540121 | POP1 | POP1 homolog, ribonuclease P/MRP subunit | Q99575 | Planar array |
| HPRR2540379 | CFAP418 | cilia and flagella associated protein 418 | Q96NL8 | Planar array |
| HPRR2540393 | TBC1D31 | TBC1 domain family member 31 | Q96DN5 | Planar array |
| HPRR2540601 | C8orf74 | chromosome 8 open reading frame 74 | Q6P047 | Planar array |
| HPRR2540604 | ESCO2 | establishment of sister chromatid cohesion N-acetyltransferase 2 | Q56NI9 | Planar array |
| HPRR2550026 | CAMK1G | calcium/calmodulin dependent protein kinase IG | Q96NX5 | Planar array |
| HPRR2550448 | KIF17 | kinesin family member 17 | Q9P2E2 | Literature |
| HPRR2550749 | CPLANE2 | ciliogenesis and Planar array polarity effector complex subunit 2 | Q9BU20 | Planar array |
| HPRR2550927 | FCN3 | ficolin 3 | O75636 | Planar array |
| HPRR2551329 | BEND5 | BEN domain containing 5 | Q7L4P6 | Planar array |
| HPRR2551394 | NOL9 | nucleolar protein 9 | Q5SY16 | Planar array |
| HPRR2551482 | BPNT1 | 3'(2'), 5'-bisphosphate nucleotidase 1 | O95861 | Planar array |
| HPRR2551635 | SPATA46 | spermatogenesis associated 46 | Q5T0L3 | Planar array |
| HPRR2570168 | HACE1 | HECT domain and ankyrin repeat containing E3 ubiquitin protein ligase 1 | Q8IYU2 | Planar array |
| HPRR2590043 | ALDH8A1 | aldehyde dehydrogenase 8 family member A1 | Q9H2A2 | Planar array |

| | | | | |
|---|---|---|---|---|
| HPRR2620090 | AFDN | afadin, adherens junction formation factor | P55196 | Planar array |
| HPRR2750059 | IGF1R | insulin like growth factor 1 receptor | P08069 | Planar array |
| HPRR2760143 | RAD23B | RAD23 homolog B, nucleotide excision repair protein | P54727 | Literature |
| HPRR2760259 | IL6R | interleukin 6 receptor | P08887 | Planar array |
| HPRR2760324 | IFNB1 | interferon beta 1 | P01574 | Literature |
| HPRR2770023 | TACR1 | tachykinin receptor 1 | P25103 | Planar array |
| HPRR2770194 | TAS2R38 | taste 2 receptor member 38 | P59533 | Planar array |
| HPRR2800140 | INPP5J | inositol polyphosphate-5-phosphatase J | Q15735 | Planar array |
| HPRR2810009 | MSL3 | MSL complex subunit 3 | Q8N5Y2 | Planar array |
| HPRR2810079 | GABRE | gamma-aminobutyric acid type A receptor subunit epsilon | P78334 | Planar array |
| HPRR2830005 | NPPB | natriuretic peptide B | P16860 | Literature |
| HPRR2850155 | FBXW4 | F-box and WD repeat domain containing 4 | P57775 | Planar array |
| HPRR2850306 | KIAA1109 | bridge-like lipid transfer protein family member 1 | Q2LD37 | Planar array |
| HPRR2850476 | ATMIN | ATM interactor | O43313 | Planar array |
| HPRR2850493 | CYP2S1 | cytochrome P450 family 2 subfamily S member 1 | Q96SQ9 | Planar array |
| HPRR2850695 | TRDN | triadin | Q13061 | Planar array |
| HPRR2920228 | COQ10B | coenzyme Q10B | Q9H8M1 | Planar array |
| HPRR2920354 | AFTPH | aftiphilin | Q6ULP2 | Planar array |
| HPRR2930211 | PID1 | phosphotyrosine interaction domain containing 1 | Q7Z2X4 | Planar array |
| HPRR2930354 | SLC4A1AP | solute carrier family 4 member 1 adaptor protein | Q9BWU0 | Planar array |
| HPRR2930445 | NCKAP5 | NCK associated protein 5 | O14513 | Planar array |
| HPRR2950061 | CERKL | ceramide kinase like | Q49MI3 | Planar array |
| HPRR2950096 | RPEL1;RPE | ribulose-5-phosphate-3-epimerase like 1;ribulose-5-phosphate-3-epimerase | Q2QD12;Q96AT9 | Planar array |
| HPRR2960293 | TAMM41 | TAM41 mitochondrial translocator assembly and maintenance homolog | Q96BW9 | Planar array |

| | | | | |
|---|---|---|---|---|
| HPRR2960328 | IQSEC1 | IQ motif and Sec7 domain ArfGEF 1 | Q6DN90 | Planar array |
| HPRR2960339 | COL8A1 | collagen type VIII alpha 1 chain | P27658 | Planar array |
| HPRR2960457 | LMOD3 | leiomodin 3 | Q0VAK6 | Planar array |
| HPRR2960533 | KIF15 | kinesin family member 15 | Q9NS87 | Literature |
| HPRR2960587 | GNL3 | G protein nucleolar 3 | Q9BVP2 | Literature |
| HPRR2960871 | METTL6 | methyltransferase 6, methylcytidine | Q8TCB7 | Planar array |
| HPRR2970026 | JAK2 | Janus kinase 2 | O60674 | Planar array |
| HPRR2970029 | RIN3 | Ras and Rab interactor 3 | Q8TB24 | Planar array |
| HPRR2990081 | CD80 | CD80 molecule | P33681 | Planar array |
| HPRR2990086 | CNTFR | ciliary neurotrophic factor receptor | P26992 | Planar array |
| HPRR3000085 | ODAM | odontogenic, ameloblast associated | A1E959 | Planar array |
| HPRR3000391 | ZFYVE28 | zinc finger FYVE-type containing 28 | Q9HCC9 | Planar array |
| HPRR3000517 | KIAA0232 | KIAA0232 | Q92628 | Planar array |
| HPRR3010180 | DMGDH | dimethylglycine dehydrogenase | Q9UI17 | Planar array |
| HPRR3010181 | BHMT2 | betaine--homocysteine S-methyltransferase 2 | Q9H2M3 | Planar array |
| HPRR3010280 | WDR36 | WD repeat domain 36 | UniProtKB unreviewed (TrEMBL) | Planar array |
| HPRR3010287 | NDUFS6 | NADH:ubiquinone oxidoreductase subunit S6 | O75380 | Planar array |
| HPRR3010342 | TENM2 | teneurin transmembrane protein 2 | Q9NT68 | Planar array |
| HPRR3010357 | SRFBP1 | serum response factor binding protein 1 | Q8NEF9 | Planar array |
| HPRR3010367 | POC5 | POC5 centriolar protein | Q8NA72 | Planar array |
| HPRR3010438 | TMEM161B | transmembrane protein 161B | Q8NDZ6 | Planar array |
| HPRR3010474 | HSPB3 | heat shock protein family B (small) member 3 | Q12988 | Planar array |
| HPRR3010494 | FAM71B | golgi associated RAB2B interactor family member 3 | Q8TC56 | Planar array |
| HPRR3010683 | TTC37 | SKI3 subunit of superkiller complex | Q6PGP7 | Planar array |

| | | | | |
|---|---|---|---|---|
| HPRR3010717 | SRA1 | steroid receptor RNA activator 1 | Q9HD15 | Planar array |
| HPRR3010723 | NEURL1B | neuralized E3 ubiquitin protein ligase 1B | A8MQ27 | Planar array |
| HPRR3020144 | MTPAP | mitochondrial poly(A) polymerase | Q9NVV4 | Planar array |
| HPRR3020186 | DNAJC12 | DnaJ heat shock protein family (Hsp40) member C12 | Q9UKB3 | Planar array |
| HPRR3020493 | ADK | adenosine kinase | P55263 | Planar array |
| HPRR3020533 | R3HCC1L | R3H domain and coiled-coil containing 1 like | Q7Z5L2 | Planar array |
| HPRR3050149 | NAA40 | N-alpha-acetyltransferase 40, NatD catalytic subunit | Q86UY6 | Planar array |
| HPRR3050173 | ELMOD1 | ELMO domain containing 1 | Q8N336 | Planar array |
| HPRR3050186 | PRPF19 | pre-mRNA processing factor 19 | Q9UMS4 | Planar array |
| HPRR3050353 | CCDC81 | coiled-coil domain containing 81 | Q6ZN84 | Planar array |
| HPRR3050471 | OOSP2 | oocyte secreted protein 2 | Q86WS3 | Planar array |
| HPRR3050498 | ARL14EP | ADP ribosylation factor like GTPase 14 effector protein | Q8N8R7 | Literature |
| HPRR3050724 | ANKRD49 | ankyrin repeat domain 49 | Q8WVL7 | Planar array |
| HPRR3050831 | EIF1AD | eukaryotic translation initiation factor 1A domain containing | Q8N9N8 | Literature |
| HPRR3050930 | C11orf54 | chromosome 11 open reading frame 54 | Q9H0W9 | Planar array |
| HPRR3060026 | TEX15 | testis expressed 15, meiosis and synapsis associated | Q9BXT5 | Planar array |
| HPRR3060143 | FBXW8 | F-box and WD repeat domain containing 8 | Q8N3Y1 | Planar array |
| HPRR3070010 | PLEKHA5 | pleckstrin homology domain containing A5 | Q9HAU0 | Planar array |
| HPRR3070018 | PLEKHG6 | pleckstrin homology and RhoGEF domain containing G6 | Q3KR16 | Planar array |
| HPRR3070036 | ANO2 | anoctamin 2 | Q9NQ90 | Planar array |
| HPRR3070082 | DIP2B | disco interacting protein 2 homolog B | Q9P265 | Planar array |
| HPRR3070159 | BIN2 | bridging integrator 2 | Q9UBW5 | Planar array |
| HPRR3070226 | PRMT8 | protein arginine methyltransferase 8 | Q9NR22 | Planar array |
| HPRR3070438 | C12orf65 | mitochondrial translation release factor in rescue | Q9H3J6 | Planar array |

| | | | | |
|---|---|---|---|---|
| HPRR3070474 | APPL2 | adaptor protein, phosphotyrosine interacting with PH domain and leucine zipper 2 | Q8NEU8 | Planar array |
| HPRR3070609 | CCDC38 | coiled-coil domain containing 38 | Q502W7 | Planar array |
| HPRR3070854 | C12orf42 | chromosome 12 open reading frame 42 | Q96LP6 | Planar array |
| HPRR3070974 | RILPL1 | Rab interacting lysosomal protein like 1 | Q5EBL4 | Planar array |
| HPRR3090326 | CCDC168 | coiled-coil domain containing 168 | Q8NDH2 | Literature |
| HPRR3100069 | NEDD4 | NEDD4 E3 ubiquitin protein ligase | P46934 | Planar array |
| HPRR3100209 | IREB2 | iron responsive element binding protein 2 | P48200 | Planar array |
| HPRR3100272 | LYSMD2 | LysM domain containing 2 | Q8IV50 | Planar array |
| HPRR3100511 | KIF7 | kinesin family member 7 | Q2M1P5 | Literature |
| HPRR3100532 | CCNDBP1 | cyclin D1 binding protein 1 | O95273 | Planar array |
| HPRR3120006 | MOCOS | molybdenum cofactor sulfurase | Q96EN8 | Planar array |
| HPRR3120222 | EMILIN2 | elastin microfibril interfacer 2 | Q9BXX0 | Planar array |
| HPRR3130014 | NEK1 | NIMA related kinase 1 | Q96PY6 | Planar array |
| HPRR3140351 | TJP3 | tight junction protein 3 | O95049 | Planar array |
| HPRR3140396 | TLX2 | T cell leukemia homeobox 2 | O43763 | Planar array |
| HPRR3140436 | IL12B | interleukin 12B | P29460 | Literature |
| HPRR3140513 | A1BG | alpha-1-B glycoprotein | P04217 | Planar array |
| HPRR3140817 | HIF3A | hypoxia inducible factor 3 subunit alpha | Q9Y2N7 | Planar array |
| HPRR3160111 | GGA2 | golgi associated, gamma adaptin ear containing, ARF binding protein 2 | Q9UJY4 | Planar array |
| HPRR3160195 | CHTF18 | chromosome transmission fidelity factor 18 | Q8WVB6 | Planar array |
| HPRR3160271 | RNF166 | ring finger protein 166 | Q96A37 | Planar array |
| HPRR3160325 | TCF25 | transcription factor 25 | Q9BQ70 | Planar array |
| HPRR3160356 | ADCY9 | adenylate cyclase 9 | O60503 | Planar array |
| HPRR3160388 | CCDC78 | coiled-coil domain containing 78 | A2IDD5 | Planar array |

| | | | | |
|---|---|---|---|---|
| HPRR3190119 | PSME3IP1 | proteasome activator subunit 3 interacting protein 1 | Q9GZU8 | Planar array |
| HPRR3190282 | MSRB1 | methionine sulfoxide reductase B1 | Q9NZV6 | Planar array |
| HPRR3200109 | CCDC74B;CCDC74A | coiled-coil domain containing 74B;coiled-coil domain containing 74A | Q96LY2;Q96AQ1 | Planar array |
| HPRR3210165 | HOOK2 | hook microtubule tethering protein 2 | Q96ED9 | Planar array |
| HPRR3210182 | POLR2E | RNA polymerase II, I and III subunit E | P19388 | Planar array |
| HPRR3250015 | GRWD1 | glutamate rich WD repeat containing 1 | Q9BQ67 | Planar array |
| HPRR3250123 | SLC8A2 | solute carrier family 8 member A2 | Q9UPR5 | Planar array |
| HPRR3250146 | COPE | COPI coat complex subunit epsilon | O14579 | Planar array |
| HPRR3250167 | THEG | theg spermatid protein | Q9P2T0 | Planar array |
| HPRR3260175 | CYP2A6;CYP2A13;CYP2A7 | cytochrome P450 family 2 subfamily A member 6;cytochrome P450 family 2 subfamily A member 13;cytochrome P450 family 2 subfamily A member 7 | P11509;Q16696;P20853 | Planar array |
| HPRR3270055 | CDKN2D | cyclin dependent kinase inhibitor 2D | P55273 | Planar array |
| HPRR3280192 | HDGFL2 | HDGF like 2 | Q7Z4V5 | Planar array |
| HPRR3280510 | PNMA8B | PNMA family member 8B | Q9ULN7 | Planar array |
| HPRR3290015 | NDUFB4 | NADH:ubiquinone oxidoreductase subunit B4 | O95168 | Planar array |
| HPRR3300056 | CPXM1 | carboxypeptidase X, M14 family member 1 | Q96SM3 | Planar array |
| HPRR3310245 | SPTLC3 | serine palmitoyltransferase long chain base subunit 3 | Q9NUV7 | Planar array |
| HPRR3320069 | PCDHB13 | protocadherin beta 13 | Q9Y5F0 | Planar array |
| HPRR3340078 | SLC25A52;SLC25A51 | solute carrier family 25 member 52;solute carrier family 25 member 51 | Q3SY17;Q9H1U9 | Planar array |
| HPRR3340100 | CCDC144A | coiled-coil domain containing 144A | A2RUR9 | Planar array |
| HPRR3340111 | FRMPD2 | FERM and PDZ domain containing 2 | Q68DX3 | Planar array |
| HPRR3340163 | PCBP1 | poly(rC) binding protein 1 | Q15365 | Planar array |
| HPRR3340233 | FKBP9 | FKBP prolyl isomerase 9 | O95302 | Planar array |

| | | | | |
|---|---|---|---|---|
| HPRR3340371 | VARS1 | valyl-tRNA synthetase 1 | P26640 | Planar array |
| HPRR3340440 | PRR20C;PRR20A;PRR20B;PRR20D;PRR20E | proline rich 20B;proline rich 20D;proline rich 20E;proline rich 20A;proline rich 20C | P86481;P86480;P86478;P86496;P86479 | Planar array |
| HPRR3360041 | IFNA16;IFNA7;IFNA10;IFNW1;IFNA21;IFNA17;IFNA14;IFNA5;IFNA2;IFNA8;IFNA1;IFNA6;IFNA13;IFNA4 | interferon alpha 16;interferon alpha 7;interferon alpha 10;interferon omega 1;interferon alpha 21;interferon alpha 17;interferon alpha 14;interferon alpha 5;interferon alpha 2;interferon alpha 8;interferon alpha 1;interferon alpha 6;interferon alpha 13;interferon alpha 4 | P05015;P01567;P01566;P05000;P01568;P01571;P01570;P01569;P01563;P32881;P01562;P05013;P05014 | Literature |
| HPRR3360076 | NAT2;NAT1 | N-acetyltransferase 2;N-acetyltransferase 1 | P11245;P18440 | Planar array |
| HPRR3400251 | DRC1 | dynein regulatory complex subunit 1 | Q96MC2 | Planar array |
| HPRR3400291 | RTP5 | receptor transporter protein 5 (putative) | Q14D33 | Planar array |
| HPRR3410115 | LYST | lysosomal trafficking regulator | Q99698 | Planar array |
| HPRR3420052 | C17orf50 | chromosome 17 open reading frame 50 | Q8WW18 | Planar array |
| HPRR3420238 | COPRS | coordinator of PRMT5 and differentiation stimulator | Q9NQ92 | Planar array |
| HPRR3420526 | KCNK4 | potassium two pore domain channel subfamily K member 4 | Q9NYG8 | Planar array |
| HPRR3420661 | ZNF669 | zinc finger protein 669 | Q96BR6 | Planar array |
| HPRR3420742 | S100A7L2 | S100 calcium-binding protein A7 | Q86SG5;P31151 | Planar array |
| HPRR3440025 | STK38 | serine/threonine kinase 38 | Q15208 | Planar array |
| HPRR3440035 | SEPTIN11 | septin 11 | Q9NVA2 | Planar array |
| HPRR3440056 | RAB3C | RAB3C, member RAS oncogene family | Q96E17 | Planar array |
| HPRR3450069 | TCF15 | transcription factor 15 | Q12870 | Planar array |
| HPRR3460051 | CRACD | capping protein inhibiting regulator of actin dynamics | Q6ZU35 | Planar array |
| HPRR3460054 | PSMC6 | proteasome 26S subunit, ATPase 6 | P62333 | Planar array |
| HPRR3460147 | USP9X | ubiquitin specific peptidase 9 X-linked | Q93008 | Planar array |
| HPRR3460778 | KRTAP17-1 | keratin associated protein 17-1 | Q9BYP8 | Planar array |
| HPRR3460790 | n.a. | novel protein | n.a. | Planar array |

| | | | | |
|---|---|---|---|---|
| HPRR3460904 | CCDC191 | coiled-coil domain containing 191 | Q8NCU4 | Planar array |
| HPRR3460919 | PDCL2 | phosducin like 2 | Q8N4E4 | Planar array |
| HPRR3470279 | PRCD | photoreceptor disc component | Q00LT1 | Planar array |
| HPRR3500307 | TPST1 | tyrosylprotein sulfotransferase 1 | O60507 | Planar array |
| HPRR3560010 | LMOD2 | leiomodin 2 | Q6P5Q4 | Planar array |
| HPRR3590069 | MRPL43 | mitochondrial ribosomal protein L43 | Q8N983 | Planar array |
| HPRR3600221 | PET117 | PET117 cytochrome c oxidase chaperone | Q6UWS5 | Planar array |
| HPRR3610061 | NFKBIB | NFKB inhibitor beta | Q15653 | Planar array |
| HPRR3610185 | C1orf53 | chromosome 1 open reading frame 53 | Q5VUE5 | Planar array |
| HPRR3620020 | OR4M1;OR4M2;OR4M2B;ENSG00000275249 | olfactory receptor family 4 subfamily M member 1;olfactory receptor family 4 subfamily M member 2;olfactory receptor family 4 subfamily M member 2B; | Q8NGD0;Q8NGB6;A0A0X1KG70 | Planar array |
| HPRR3670014 | PATZ1 | POZ/BTB and AT hook containing zinc finger 1 | Q9HBE1 | Planar array |
| HPRR3680006 | SPACA6 | sperm acrosome associated 6 | W5XKT8 | Planar array |
| HPRR3690140 | OR2Z1 | olfactory receptor family 2 subfamily Z member 1 | Q8NG97 | Planar array |
| HPRR3700073 | ZBTB25 | zinc finger and BTB domain containing 25 | P24278 | Planar array |
| HPRR3700118 | TGFB1 | transforming growth factor beta 1 | B2C8Z3 | Literature |
| HPRR3700165 | NID1 | nidogen 1 | P14543 | Planar array |
| HPRR3700438 | C16orf74 | chromosome 16 open reading frame 74 | Q96GX8 | Planar array |
| HPRR3710011 | KLLN | killin, p53 regulated DNA replication inhibitor | B2CW77 | Planar array |
| HPRR3720135 | ADGRF5 | adhesion G protein-coupled receptor F5 | Q8IZF2 | Planar array |
| HPRR3720275 | CTTN | cortactin | Q14247 | Planar array |
| HPRR3720532 | SNAPC2 | small nuclear RNA activating complex polypeptide 2 | Q13487 | Literature |
| HPRR3720670 | EIF3B | eukaryotic translation initiation factor 3 subunit B | P55884 | Planar array |
| HPRR3730069 | IFNG | interferon gamma | P01579 | Literature |

| | | | | |
|---|---|---|---|---|
| HPRR3730126 | VEGFA | vascular endothelial growth factor A | P15692 | Literature |
| HPRR3730160 | SLC4A9 | solute carrier family 4 member 9 | Q96Q91 | Planar array |
| HPRR3730226 | INVS | inversin | Q9Y283 | Planar array |
| HPRR3760008 | IGFBPL1 | insulin like growth factor binding protein like 1 | Q8WX77 | Planar array |
| HPRR3760127 | NCOA2 | nuclear receptor coactivator 2 | Q15596 | Planar array |
| HPRR3760231 | SLC24A4 | solute carrier family 24 member 4 | Q8NFF2 | Planar array |
| HPRR3760489 | KCNJ6 | potassium inwardly rectifying channel subfamily J member 6 | P48051 | Planar array |
| HPRR3760628 | AGPAT3 | 1-acylglycerol-3-phosphate O-acyltransferase 3 | Q9NRZ7 | Planar array |
| HPRR3760680 | ZNF283 | zinc finger protein 283 | Q8N7M2 | Planar array |
| HPRR3760694 | EMC10 | ER membrane protein complex subunit 10 | Q5UCC4 | Planar array |
| HPRR3761134 | TM4SF20 | transmembrane 4 L six family member 20 | Q53R12 | Planar array |
| HPRR3770042 | PTPRS | protein tyrosine phosphatase receptor type S | Q13332 | Planar array |
| HPRR3780091 | C1QB | complement C1q B chain | P02746 | Literature |
| HPRR3790073 | LCN15 | lipocalin 15 | Q6UWW0 | Planar array |
| HPRR3790222 | RNASE10 | ribonuclease A family member 10 (inactive) | Q5GAN6 | Planar array |
| HPRR3790443 | ZNF699 | zinc finger protein 699 | Q32M78 | Planar array |
| HPRR3790485 | SLC18A3 | solute carrier family 18 member A3 | Q16572 | Planar array |
| HPRR3790901 | TAS2R41 | taste 2 receptor member 41 | P59536 | Planar array |
| HPRR3830040 | MAZ | MYC associated zinc finger protein | P56270 | Planar array |
| HPRR3830348 | LKAAEAR1 | LKAAEAR motif containing 1 | Q8TD35 | Planar array |
| HPRR3830412 | TRHR | thyrotropin releasing hormone receptor | P34981 | Planar array |
| HPRR3830590 | FZD9 | frizzled class receptor 9 | O00144 | Planar array |
| HPRR3830598 | CEBPA | CCAAT enhancer binding protein alpha | P49715 | Literature |
| HPRR3860150 | FAM120A | family with sequence similarity 120A | Q9NZB2 | Planar array |
| HPRR3860330 | KCNQ2 | potassium voltage-gated channel subfamily Q member 2 | O43526 | Planar array |

| | | | | |
|---|---|---|---|---|
| HPRR3870206 | TCFL5 | transcription factor like 5 | Q9UL49 | Planar array |
| HPRR3870291 | HCFC1R1 | host cell factor C1 regulator 1 | Q9NWW0 | Planar array |
| HPRR3890079 | TBP | TATA-box binding protein | P20226 | Planar array |
| HPRR3890290 | KDM3B | lysine demethylase 3B | Q7LBC6 | Planar array |
| HPRR3890580 | CLIP1 | CAP-Gly domain containing linker protein 1 | P30622 | Planar array |
| HPRR3890692 | ECHDC3 | enoyl-CoA hydratase domain containing 3 | Q96DC8 | Planar array |
| HPRR3890759 | AGTPBP1 | ATP/GTP binding carboxypeptidase 1 | Q9UPW5 | Planar array |
| HPRR3890914 | BCAR3 | BCAR3 adaptor protein, NSP family member | O75815 | Planar array |
| HPRR3910089 | SNORC(C2orf82) | secondary ossification center associated regulator of chondrocyte maturation | Q6UX34 | Planar array |
| HPRR3920257 | MRPS5 | mitochondrial ribosomal protein S5 | P82675 | Planar array |
| HPRR3920292 | ATL3 | atlastin GTPase 3 | Q6DD88 | Planar array |
| HPRR3950245 | BATF | basic leucine zipper ATF-like transcription factor | Q16520 | Planar array |
| HPRR3970036 | RPL29 | ribosomal protein L29 | P47914 | Planar array |
| HPRR3990019 | ATXN7L3B | ataxin 7 like 3B | Q96GX2 | Planar array |
| HPRR4000010 | BMERB1 | bMERB domain containing 1 | Q96MC5 | Planar array |
| HPRR4000037 | RIMKLB | ribosomal modification protein rimK like family member B | Q9ULI2 | Planar array |
| HPRR4000254 | MFSD2A | major facilitator superfamily domain containing 2A | Q8NA29 | Planar array |
| HPRR4030059 | ANOS1 | anosmin 1 | P23352 | Literature |
| HPRR4030078 | SMARCD3 | SWI/SNF related, matrix associated, actin dependent regulator of chromatin, subfamily d, member 3 | Q6STE5 | Planar array |
| HPRR4030107 | IGHMBP2 | immunoglobulin mu DNA binding protein 2 | P38935 | Planar array |
| HPRR4040205 | VEGFB | vascular endothelial growth factor B | P49765 | Literature |
| HPRR4040340 | TEX36 | testis expressed 36 | Q5VZQ5 | Planar array |
| HPRR4040612 | MEX3C | mex-3 RNA binding family member C | Q5U5Q3 | Planar array |

| | | | | |
|---|---|---|---|---|
| HPRR4050483 | ABCB8 | ATP binding cassette subfamily B member 8 | Q9NUT2 | Planar array |
| HPRR4050658 | SLFN12L | schlafen family member 12 like | Q6IEE8 | Planar array |
| HPRR4050663 | CCDC85C | coiled-coil domain containing 85C | A6NKD9 | Planar array |
| HPRR4060148 | ZBTB45 | zinc finger and BTB domain containing 45 | Q96K62 | Literature |
| HPRR4070046 | VSTM5 | V-set and transmembrane domain containing 5 | A8MXK1 | Planar array |
| HPRR4080071 | RHOG | ras homolog family member G | P84095 | Literature |
| HPRR4110390 | NPAS1 | neuronal PAS domain protein 1 | Q99742 | Planar array |
| HPRR4120003 | TMEM123 | transmembrane protein 123 | Q8N131 | Planar array |
| HPRR4120010 | B4GALT5 | beta-1,4-galactosyltransferase 5 | O43286 | Planar array |
| HPRR4120192 | RMI2 | RecQ mediated genome instability 2 | Q96E14 | Planar array |
| HPRR4160013 | ZNF195 | zinc finger protein 195 | O14628 | Planar array |
| HPRR4160919 | PPP4R3A | protein phosphatase 4 regulatory subunit 3A | Q6IN85 | Planar array |
| HPRR4170045 | OAZ1 | ornithine decarboxylase antizyme 1 | P54368 | Planar array |
| HPRR4180321 | LIFR | LIF receptor subunit alpha | P42702 | Planar array |
| HPRR4180322 | ABCC5 | ATP binding cassette subfamily C member 5 | O15440 | Planar array |
| HPRR4180371 | SMG7 | SMG7 nonsense mediated mRNA decay factor | Q92540 | Planar array |
| HPRR4180489 | ZSCAN18 | zinc finger and SCAN domain containing 18 | Q8TBC5 | Planar array |
| HPRR4180530 | ZMIZ2 | zinc finger MIZ-type containing 2 | Q8NF64 | Planar array |
| HPRR4180650 | DDX54 | DEAD-box helicase 54 | Q8TDD1 | Planar array |
| HPRR4180663 | SYT5 | synaptotagmin 5 | O00445 | Planar array |
| HPRR4180820 | CALU | calumenin | O43852 | Planar array |
| HPRR4180933 | MICAL2 | microtubule associated monooxygenase, calponin and LIM domain containing 2 | O94851 | Planar array |
| HPRR4181020 | TBX3 | T-box transcription factor 3 | O15119 | Planar array |
| HPRR4190245 | CDCA7 | cell division cycle associated 7 | Q9BWT1 | Planar array |

| | | | | |
|---|---|---|---|---|
| HPRR4190823 | ATF3 | activating transcription factor 3 | P18847 | Planar array |
| HPRR4190832 | ANKLE1 | ankyrin repeat and LEM domain containing 1 | Q8NAG6 | Planar array |
| HPRR4200044 | MDM2 | MDM2 proto-oncogene | Q00987 | Planar array |
| HPRR4200073 | IL27RA | interleukin 27 receptor subunit alpha | Q6UWB1 | Planar array |
| HPRR4200133 | FXYD2 | FXYD domain containing ion transport regulator 2 | P54710 | Planar array |
| HPRR4220244 | NSD1 | nuclear receptor binding SET domain protein 1 | Q96L73 | Planar array |
| HPRR4220339 | CCDC88B | coiled-coil domain containing 88B | A6NC98 | Planar array |
| HPRR4220456 | FSHR | follicle stimulating hormone receptor | P23945 | Planar array |
| HPRR4240042 | UBE2O | ubiquitin conjugating enzyme E2 O | Q9C0C9 | Planar array |
| HPRR4280073 | ATP5MC1 | ATP synthase membrane subunit c locus 1 | P05496 | Planar array |
| HPRR4280075 | SOX15 | SRY-box transcription factor 15 | O60248 | Planar array |
| HPRR4280108 | AVPR1A | arginine vasopressin receptor 1A | P37288 | Literature |
| HPRR4290879 | FOXJ3 | forkhead box J3 | Q9UPW0 | Planar array |
| HPRR4320135 | NCAPH2 | non-SMC condensin II complex subunit H2 | Q6IBW4 | Planar array |
| HPRR4320502 | PLPPR4 | phospholipid phosphatase related 4 | Q7Z2D5 | Planar array |
| HPRR4340053 | CDC42EP1 | CDC42 effector protein 1 | Q00587 | Planar array |
| HPRR4340274 | EPHA1 | EPH receptor A1 | P21709 | Planar array |
| HPRR4350098 | FAM114A1 | family with sequence similarity 114 member A1 | Q8IWE2 | Planar array |
| HPRR4390250 | ESRRA | estrogen related receptor alpha | P11474 | Planar array |
| HPRR4430241 | TGFBR3L | transforming growth factor beta receptor 3 like | H3BV60 | Planar array |
| HPRR4450262 | CLDN2 | claudin 2 | P57739 | Planar array |
| HPRR4470247 | HLA-F;HLA-E;HLA-C;HLA-G;HLA-B;HLA-A | major histocompatibility complex, class I, F;major histocompatibility complex, class I, E;major histocompatibility complex, class I, C;major histocompatibility complex, class I, G;major histocompatibility complex, class I, | P30511;P13747;P10321;P17693;P01889;P04439 | Literature |

| | | | | |
|---|---|---|---|---|
| | | B;major histocompatibility complex, class I, A | | |
| HPRR4520067 | FCN2 | ficolin 2 | Q15485 | Literature |
| HPRR330265 | IL23A | interleukin 23 subunit alpha | Q9NPF7 | Literature |
| HPRR141227 | HIF1A | hypoxia inducible factor 1 subunit alpha | Q16665 | Literature |
| HPRR260285 | STAT3 | signal transducer and activator of transcription 3 | P40763 | Literature |
| HPRR330193 | PROC | protein C, inactivator of coagulation factors Va and VIIIa | P04070 | Planar array |
| HPRR330201 | ICAM1 | intercellular adhesion molecule 1 | P05362 | Literature |
| HPRR1030014 | MUC7 | mucin 7, secreted | Q8TAX7 | Planar array |
| HPRR1950251 | CFAP61 | cilia and flagella associated protein 61 | Q8NHU2 | Planar array |
| HPRR1950277 | CDHR5 | cadherin related family member 5 | Q9HBB8 | Planar array |
| HPRR2140286 | CALHM3 | calcium homeostasis modulator 3 | Q86XJ0 | Planar array |
| HPRR3860359 | IGF2BP2 | insulin like growth factor 2 mRNA binding protein 2 | Q9Y6M1 | Planar array |
| HPRR4220019 | RIGI(DDX58) | RNA sensor RIG-I(DEAD Box Protein 58) | O95786 | Planar array |
| HPRR4220163 | AMOTL1 | angiomotin like 1 | Q8IY63 | Planar array |
| HPRR140840 | SERPINA6 | serpin family A member 6 | P08185 | Literature |
| HPRR3070561 | LMNTD1 | lamin tail domain containing 1 | Q8N9Z9 | Planar array |
| HPRR3400036 | STARD7 | StAR related lipid transfer domain containing 7 | Q9NQZ5 | Planar array |
| HPRR1420001 | SELE | selectin E | P16581 | Literature |
| HPRR2550020 | SELE | selectin E | P16581 | Literature |
| HPRR2740005 | KIF4A;KIF4B | kinesin family member 4A;kinesin family member 4B | O95239;Q2VIQ3 | Literature |
| HPRR2740007 | KIF4A;KIF4B | kinesin family member 4A;kinesin family member 4B | O95239;Q2VIQ3 | Literature |
| HPRR3070561 | LMNTD1 | lamin tail domain containing 1 | Q8N9Z9 | Planar array |
| HPRR3400036 | STARD7 | StAR related lipid transfer domain containing 7 | Q9NQZ5 | Planar array |
| HPRR3450239 | TNF | tumor necrosis factor | P01375 | Literature |
| HPRR3450240 | TNF | tumor necrosis factor | P01375 | Literature |
| HPRR4390019 | SERPINA6 | serpin family A member 6 | P08185 | Literature |

| | | | | |
|---|---|---|---|---|
| HPRR260123 | APOA4 | apolipoprotein A4 | P06727 | Literature |
| HPRR260124 | APOA4 | apolipoprotein A4 | P06727 | Literature |
| HPRR320020 | SERPINA1 | serpin family A member 1 | P01009 | Literature |
| HPRR320021 | SERPINA1 | serpin family A member 1 | P01009 | Literature |

**Supplementary Table S3. Full cohort targeted screening. Antigens´ information.**

| Antigen ID | Gene name | Gene Description | Uniprot ID | Inclusion criteria |
|---|---|---|---|---|
| HPRR140351 | EDDM3B | epididymal protein 3B | P56851 | Pilot screening |
| HPRR141227 | HIF1A | hypoxia inducible factor 1 subunit alpha | Q16665 | Pilot screening |
| HPRR232315 | LAMP2 | lysosomal associated membrane protein 2 | P13473 | Literature |
| HPRR252447 | NLRP3 | NLR family pyrin domain containing 3 | Q96P20 | Literature |
| HPRR260123 | APOA4 | apolipoprotein A4 | P06727 | Pilot screening |
| HPRR260245 | CD36 | CD36 molecule | P16671 | Literature |
| HPRR280090 | C3 | complement C3 | P01024 | Pilot screening |
| HPRR280093 | C5 | complement C5 | P01031 | Literature |
| HPRR330007 | IL6 | interleukin 6 | P05231 | Pilot screening |
| HPRR330039 | IFNG | interferon gamma | P01579 | Literature |
| HPRR330196 | THBD | thrombomodulin | P07204 | Literature |
| HPRR330199 | SELP | selectin P | P16109 | Literature |
| HPRR330201 | ICAM1 | intercellular adhesion molecule 1 | P05362 | Pilot screening |
| HPRR330203 | ICAM2 | intercellular adhesion molecule 2 | P13598 | Literature |
| HPRR330265 | IL23A | interleukin 23 subunit alpha | Q9NPF7 | Pilot screening |
| HPRR330279 | IL1RN | interleukin 1 receptor antagonist | P18510 | Literature |
| HPRR350154 | VCAM1 | vascular cell adhesion molecule 1 | P19320 | Literature |
| HPRR360310 | SIM2 | SIM bHLH transcription factor 2 | Q14190 | Literature |
| HPRR610027 | CXCL9 | C-X-C motif chemokine ligand 9 | Q07325 | Literature |
| HPRR610046 | CCL2 | C-C motif chemokine ligand 2 | P13500 | Literature |
| HPRR670018 | PLAUR | plasminogen activator, urokinase receptor | Q03405 | Literature |
| HPRR670290 | PECAM1 | platelet and endothelial cell adhesion molecule 1 | P16284 | Literature |
| HPRR1030014 | MUC7 | mucin 7, secreted | Q8TAX7 | Pilot screening |
| HPRR1060025 | C3AR1 | complement C3a receptor 1 | Q16581 | Literature |
| HPRR1310105 | ITGB2 | integrin subunit beta 2 | P05107 | Literature |
| HPRR1390031 | SPP1 | secreted phosphoprotein 1 | P10451 | Literature |
| HPRR1420001 | SELE | selectin E | P16581 | Pilot screening |

| | | | | |
|---|---|---|---|---|
| HPRR1450742 | RBPJ | recombination signal binding protein for immunoglobulin kappa J region | Q06330 | Pilot screening |
| HPRR1480001 | RAB7A | RAB7A, member RAS oncogene family | P51149 | Literature |
| HPRR1820001 | CCL18 | C-C motif chemokine ligand 18 | P55774 | Literature |
| HPRR1850063 | ALDH1A2 | aldehyde dehydrogenase 1 family member A2 | O94788 | Pilot screening |
| HPRR1920067 | NAPG | NSF attachment protein gamma | Q99747 | Pilot screening |
| HPRR1950036 | HFE | homeostatic iron regulator | Q30201 | Pilot screening |
| HPRR1950558 | IL1RL1 | interleukin 1 receptor like 1 | Q01638 | Literature |
| HPRR1950559 | IL1RL1 | interleukin 1 receptor like 1 | Q01638 | Literature |
| HPRR1950646 | LRP11 | LDL receptor related protein 11 | Q86VZ4 | Pilot screening |
| HPRR1950853 | KLB | klotho beta | Q86Z14 | Pilot screening |
| HPRR1951629 | CXCL11 | C-X-C motif chemokine ligand 11 | O14625 | Literature |
| HPRR2050262 | ENTPD1 | ectonucleoside triphosphate diphosphohydrolase 1 | P49961 | Literature |
| HPRR2090126 | EDNRA | endothelin receptor type A | P25101 | Literature |
| HPRR2090127 | EDNRA | endothelin receptor type A | P25101 | Literature |
| HPRR2150261 | CLDN18 | claudin 18 | P56856 | Pilot screening |
| HPRR2260138 | UBQLN3 | ubiquilin 3 | Q9H347 | Pilot screening |
| HPRR2310082 | GDF3 | growth differentiation factor 3 | Q9NR23 | Pilot screening |
| HPRR2370088 | ADAMTS18 | ADAM metallopeptidase with thrombospondin type 1 motif 18 | Q8TE60 | Literature |
| HPRR2460064 | PDE2A | phosphodiesterase 2A | O00408 | Pilot screening |
| HPRR2470419 | WDR31 | WD repeat domain 31 | Q8NA23 | Pilot screening |
| HPRR2550026 | CAMK1G | calcium/calmodulin dependent protein kinase IG | Q96NX5 | Pilot screening |
| HPRR2550099 | KIF1B | kinesin family member 1B | O60333 | Literature |
| HPRR2550100 | KIF1B | kinesin family member 1B | O60333 | Literature |
| HPRR2550749 | CPLANE2 | ciliogenesis and Planar array polarity effector 2 | Q9BU20 | Pilot screening |

| | | | | |
|---|---|---|---|---|
| HPRR2550927 | FCN3 | ficolin 3 | O75636 | Pilot screening |
| HPRR2590043 | ALDH8A1 | aldehyde dehydrogenase 8 family member A1 | Q9H2A2 | Pilot screening |
| HPRR2700191 | CXCL10 | C-X-C motif chemokine ligand 10 | P02778 | Literature |
| HPRR2700195 | CXCL8 | C-X-C motif chemokine ligand 8 | P10145 | Literature |
| HPRR2740005 | KIF4A;KIF4B | kinesin family member 4A;kinesin family member 4B | O95239;Q2VIQ3 | Pilot screening |
| HPRR2740007 | KIF4A;KIF4B | kinesin family member 4A;kinesin family member 4B | O95239;Q2VIQ3 | Pilot screening |
| HPRR2760140 | SPP1 | secreted phosphoprotein 1 | P10451 | Literature |
| HPRR2810012 | LAMP2 | lysosomal associated membrane protein 2 | P13473 | Literature |
| HPRR2810013 | LAMP2 | lysosomal associated membrane protein 2 | P13473 | Literature |
| HPRR2850155 | FBXW4 | F-box and WD repeat domain containing 4 | P57775 | Pilot screening |
| HPRR2930359 | KIF5C | kinesin family member 5C | O60282 | Literature |
| HPRR2960293 | TAMM41 | TAM41 mitochondrial translocator assembly and maintenance homolog | Q96BW9 | Pilot screening |
| HPRR2960531 | KIF15 | kinesin family member 15 | Q9NS87 | Pilot screening |
| HPRR2960533 | KIF15 | kinesin family member 15 | Q9NS87 | Pilot screening |
| HPRR2960871 | METTL6 | methyltransferase like 6 | Q8TCB7 | Pilot screening |
| HPRR2990081 | CD80 | CD80 molecule | P33681 | Pilot screening |
| HPRR3000085 | ODAM | odontogenic, ameloblast associated | A1E959 | Pilot screening |
| HPRR3000391 | ZFYVE28 | zinc finger FYVE-type containing 28 | Q9HCC9 | Pilot screening |
| HPRR3010367 | POC5 | POC5 centriolar protein | Q8NA72 | Pilot screening |
| HPRR3010494 | FAM71B | family with sequence similarity 71 member B | Q8TC56 | Pilot screening |
| HPRR3050498 | ARL14EP | ADP ribosylation factor like GTPase 14 effector protein | Q8N8R7 | Pilot screening |
| HPRR3050931 | C11orf54;AP001273.2 | chromosome 11 open reading frame 54;novel protein, C11orf54-MED17 readthrough | Q9H0W9 | Pilot screening |
| HPRR3070561 | LMNTD1 | lamin tail domain containing 1 | Q8N9Z9 | Pilot screening |
| HPRR3100272 | LYSMD2 | LysM domain containing 2 | Q8IV50 | Pilot screening |

| | | | | |
|---|---|---|---|---|
| HPRR3120006 | MOCOS | molybdenum cofactor sulfurase | Q96EN8 | Pilot screening |
| HPRR3140396 | TLX2 | T cell leukemia homeobox 2 | O43763 | Pilot screening |
| HPRR3140639 | IL2RA | interleukin 2 receptor subunit alpha | P01589 | Literature |
| HPRR3210165 | HOOK2 | hook microtubule tethering protein 2 | Q96ED9 | Pilot screening |
| HPRR3340371 | VARS1 | valyl-tRNA synthetase 1 | P26640 | Pilot screening |
| HPRR3420052 | C17orf50 | chromosome 17 open reading frame 50 | Q8WW18 | Pilot screening |
| HPRR3420526 | KCNK4 | potassium two pore domain channel subfamily K member 4 | Q9NYG8 | Pilot screening |
| HPRR3700150 | TIMP1 | TIMP metallopeptidase inhibitor 1 | P01033 | Literature |
| HPRR3700165 | NID1 | nidogen 1 | P14543 | Pilot screening |
| HPRR3700438 | C16orf74 | chromosome 16 open reading frame 74 | Q96GX8 | Pilot screening |
| HPRR3760008 | IGFBPL1 | insulin like growth factor binding protein like 1 | Q8WX77 | Pilot screening |
| HPRR3760412 | ITGB1 | integrin subunit beta 1 | P05556 | Literature |
| HPRR3860330 | KCNQ2 | potassium voltage-gated channel subfamily Q member 2 | O43526 | Pilot screening |
| HPRR3860359 | IGF2BP2 | insulin like growth factor 2 mRNA binding protein 2 | Q9Y6M1 | Pilot screening |
| HPRR3880292 | AC099811.2;RAB5C;RAB5B;RAB5A | novel protein;RAB5C, member RAS oncogene family;RAB5B, member RAS oncogene family;RAB5A, member RAS oncogene family | P51148;P61020;P20339 | Literature |
| HPRR3900013 | MTOR | mechanistic target of rapamycin kinase | P42345 | Literature |
| HPRR3920084 | KRAS | KRAS proto-oncogene, GTPase | P01116 | Literature |
| HPRR3920257 | MRPS5 | mitochondrial ribosomal protein S5 | P82675 | Pilot screening |
| HPRR3920292 | ATL3 | atlastin GTPase 3 | Q6DD88 | Pilot screening |
| HPRR3990019 | ATXN7L3B | ataxin 7 like 3B | Q96GX2 | Pilot screening |
| HPRR4000010 | BMERB1 | bMERB domain containing 1 | Q96MC5 | Pilot screening |
| HPRR4000320 | KIF5C | kinesin family member 5C | O60282 | Literature |
| HPRR4030067 | KITLG | KIT ligand | P21583 | Literature |

| | | | | |
|---|---|---|---|---|
| HPRR4120003 | TMEM123 | transmembrane protein 123 | Q8N131 | Pilot screening |
| HPRR4120192 | RMI2 | RecQ mediated genome instability 2 | Q96E14 | Pilot screening |
| HPRR4160673 | FLT3LG | fms related receptor tyrosine kinase 3 ligand | P49771 | Literature |
| HPRR4170045 | OAZ1 | ornithine decarboxylase antizyme 1 | P54368 | Pilot screening |
| HPRR4180530 | ZMIZ2 | zinc finger MIZ-type containing 2 | Q8NF64 | Pilot screening |
| HPRR4180650 | DDX54 | DEAD-box helicase 54 | Q8TDD1 | Pilot screening |
| HPRR4180663 | SYT5 | synaptotagmin 5 | O00445 | Pilot screening |
| HPRR4180933 | MICAL2 | microtubule associated monooxygenase, calponin and LIM domain containing 2 | O94851 | Pilot screening |
| HPRR4181020 | TBX3 | T-box transcription factor 3 | O15119 | Pilot screening |
| HPRR4190823 | ATF3 | activating transcription factor 3 | P18847 | Pilot screening |
| HPRR4190832 | ANKLE1 | ankyrin repeat and LEM domain containing 1 | Q8NAG6 | Pilot screening |
| HPRR4200073 | IL27RA | interleukin 27 receptor subunit alpha | Q6UWB1 | Pilot screening |
| HPRR4200133 | FXYD2 | FXYD domain containing ion transport regulator 2 | P54710 | Pilot screening |
| HPRR4240042 | UBE2O | ubiquitin conjugating enzyme E2 O | Q9C0C9 | Pilot screening |
| HPRR4280073 | ATP5MC1 | ATP synthase membrane subunit c locus 1 | P05496 | Pilot screening |
| HPRR4280112 | PTX3 | pentraxin 3 | P26022 | Literature |
| HPRR4290799 | RAB11B;RAB11A | RAB11B, member RAS oncogene family;RAB11A, member RAS oncogene family | Q15907;P62491 | Literature |
| HPRR4290879 | FOXJ3 | forkhead box J3 | Q9UPW0 | Pilot screening |
| HPRR4320502 | PLPPR4 | phospholipid phosphatase related 4 | Q7Z2D5 | Pilot screening |
| HPRR4320529 | C5 | complement C5 | P01031 | Literature |
| HPRR4340053 | CDC42EP1 | CDC42 effector protein 1 | Q00587 | Pilot screening |
| HPRR4370105 | IL1B | interleukin 1 beta | P01584 | Literature |
| HPRR4390019 | SERPINA6 | serpin family A member 6 | P08185 | Pilot screening |
| HPRR4430241 | TGFBR3L | transforming growth factor beta receptor 3 like | H3BV60 | Pilot screening |

| | | | | |
|---|---|---|---|---|
| HPRR4520067 | FCN2 | ficolin 2 | Q15485 | Pilot screening |

**Supplementary Table S4. Baseline features included in the regression models.**

| | **Feature*** |
|---|---|
| 1 | Sex |
| 2 | Vital status |
| 3 | ANCA |
| 4 | Age |
| 5 | Main diagnosis |
| 6 | Kidney |
| 7 | Eye |
| 8 | Lung granuloma |
| 9 | Lung haemorrage |
| 10 | Interstitial lung disease |
| 11 | Ear-nose-throat |
| 12 | Trachea |
| 13 | Mucocutaneous |
| 14 | Muscoskeletal |
| 15 | Central nervous system |
| 16 | Peripheral nervous system |
| 17 | Abdominal |
| 18 | Cardiovascular |
| 19 | Other organ |

*Features 6-19 refer to affected organs

**Supplementary Table S5. Regression models information.**

| Model | Antibody Type | Features | Penalty | Outcome |
|---|---|---|---|---|
| Model 1 | Binary | Antibodies | Antibodies | Binary (Relapse/no Relapse) |
| Model 2 | Binary | Antibodies + Baseline features (continuous, categorical) | Antibodies + Baseline features | Binary (Relapse/no Relapse) |
| Model 3 | Binary | Antibodies + Baseline features (continuous, categorical) | Antibodies only | Binary (Relapse/no Relapse) |
| Model 4 | Binary | Antibodies | Antibodies | Continuous (Relapse per Year) |
| Model 5 | Binary | Antibodies + Baseline features (continuous, categorical) | Antibodies + Baseline features | Continuous (Relapse per Year) |
| Model 6 | Binary | Antibodies + Baseline features (continuous, categorical) | Antibodies only | Continuous (Relapse per Year) |
| Model 7 | Binary | Antibodies | Antibodies | Time to event (Time to first relapse after sample collection date |
| Model 8 | Binary | Antibodies + Baseline features (continuous, categorical) | Antibodies + Baseline features | Time to event (Time to first relapse after sample collection date |
| Model 9 | Binary | Antibodies + Baseline features (continuous, categorical) | Antibodies only | Time to event (Time to first relapse after sample collection date |
| Model 10 | Continuous | Antibodies | Antibodies | Binary (Relapse/no Relapse) |
| Model 11 | Continuous | Antibodies + Baseline features (continuous, categorical) | Antibodies + Baseline features | Binary (Relapse/no Relapse) |
| Model 12 | Continuous | Antibodies + Baseline features (continuous, categorical) | Antibodies only | Binary (Relapse/no Relapse) |
| Model 13 | Continuous | Antibodies | Antibodies | Continuous (Relapse per Year) |
| Model 14 | Continuous | Antibodies + Baseline features (continuous, categorical) | Antibodies + Baseline features | Continuous (Relapse per Year) |
| Model 15 | Continuous | Antibodies + Baseline features (continuous, categorical) | Antibodies only | Continuous (Relapse per Year) |

| | | | | |
|---|---|---|---|---|
| Model 16 | Continuous | Antibodies | Antibodies | Time to event<br>(Time to first relapse after sample collection date |
| Model 17 | Continuous | Antibodies +<br>Baseline features (continuous, categorical) | Antibodies + Baseline features | Time to event<br>(Time to first relapse after sample collection date |
| Model 18 | Continuous | Antibodies +<br>Baseline features (continuous, categorical) | Antibodies only | Time to event<br>(Time to first relapse after sample collection date |
| Model 19 | Normalized | Antibodies | Antibodies | Binary<br>(Relapse/no Relapse) |
| Model 20 | Normalized | Antibodies +<br>Baseline features (continuous, categorical) | Antibodies + Baseline features | Binary<br>(Relapse/no Relapse) |
| Model 21 | Normalized | Antibodies +<br>Baseline features (continuous, categorical) | Antibodies only | Binary<br>(Relapse/no Relapse) |
| Model 22 | Normalized | Antibodies | Antibodies | Continuous<br>(Relapse per Year) |
| Model 23 | Normalized | Antibodies +<br>Baseline features (continuous, categorical) | Antibodies + Baseline features | Continuous<br>(Relapse per Year) |
| Model 24 | Normalized | Antibodies +<br>Baseline features (continuous, categorical) | Antibodies only | Continuous<br>(Relapse per Year) |
| Model 25 | Normalized | Antibodies | Antibodies | Time to event<br>(Time to first relapse after sample collection date |
| Model 26 | Normalized | Antibodies +<br>Baseline features (continuous, categorical) | Antibodies + Baseline features | Time to event<br>(Time to first relapse after sample collection date |
| Model 27 | Normalized | Antibodies +<br>Baseline features (continuous, categorical) | Antibodies only | Time to event<br>(Time to first relapse after sample collection date |

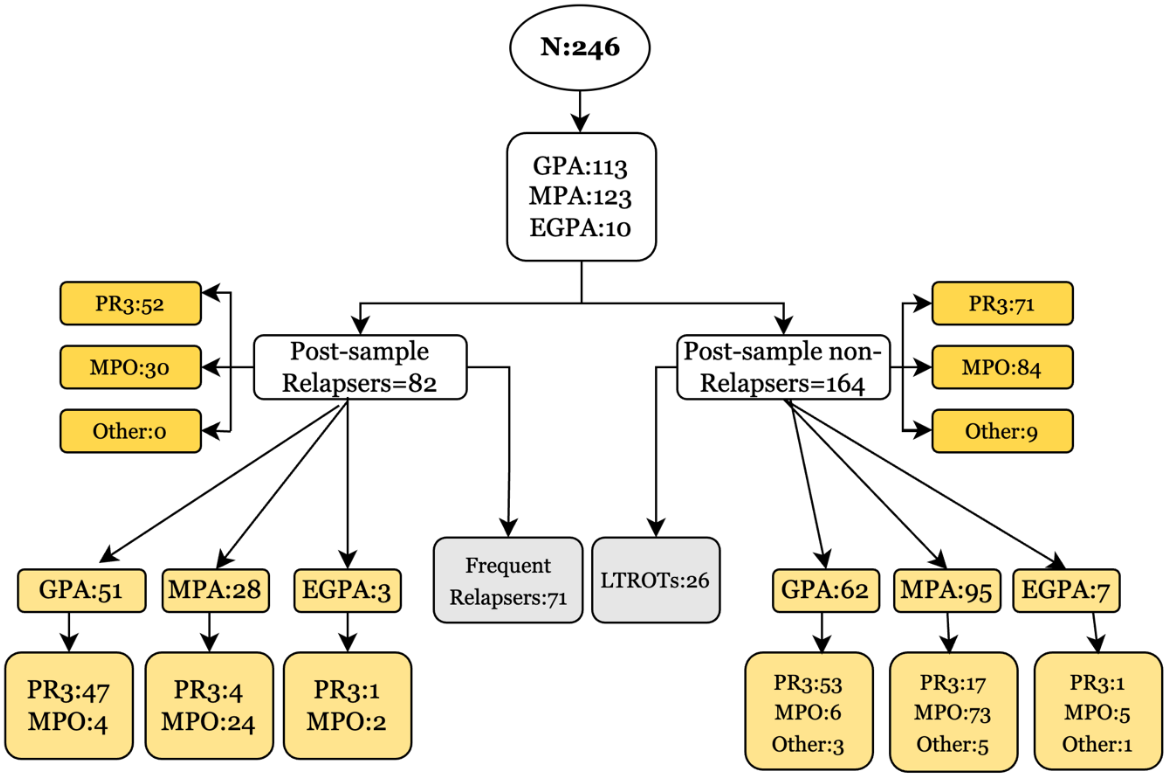


**Supplementary Figure S1**. **Classification of patients included in the study.** The flowchart represents the subclassification of the patients included in the study based on diagnosis, ANCA serology and their status in terms of relapse.

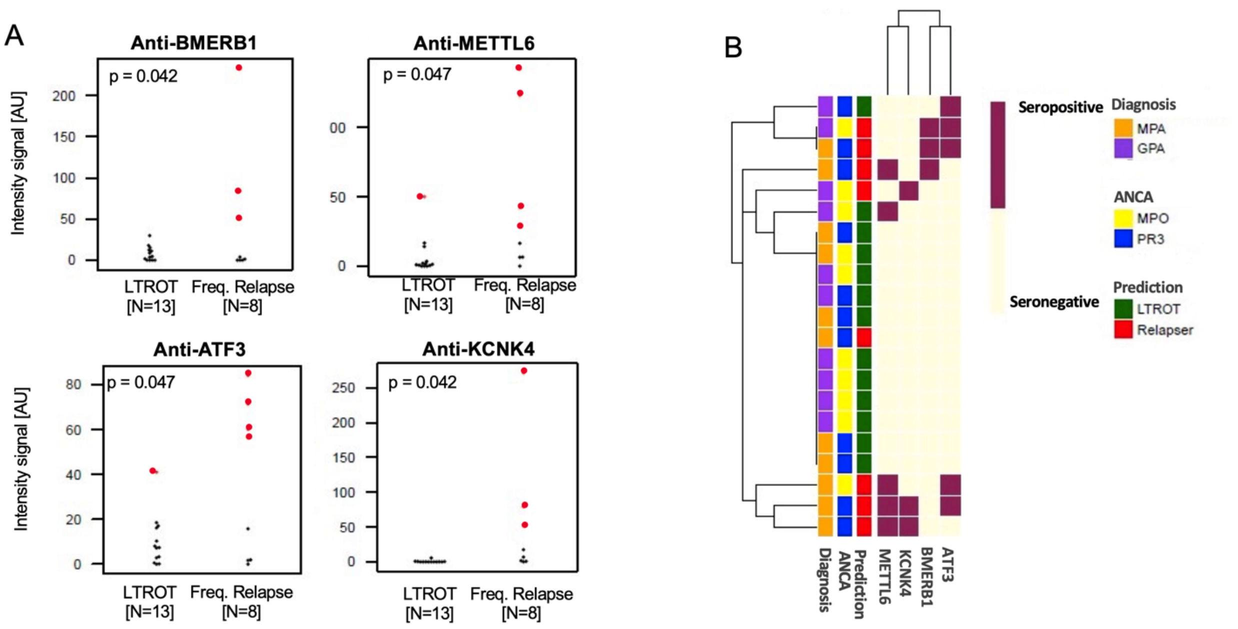


**Supplementary Figure S2. Pilot targeted screening: protein fragment specific autoantibodies.** A) Four autoantibodies were found to be more prevalent in the AAV frequent relapsers group compared to LTROT patients. Each dot in the plot represents one individual, and red dots represent individuals where the signal passed the cutoff of reactivity. P-values refer to Fisher´s exact test. B) Heatmap showing the distribution of seropositivity to the 4 autoantibodies reported in A across the 21 tested samples.

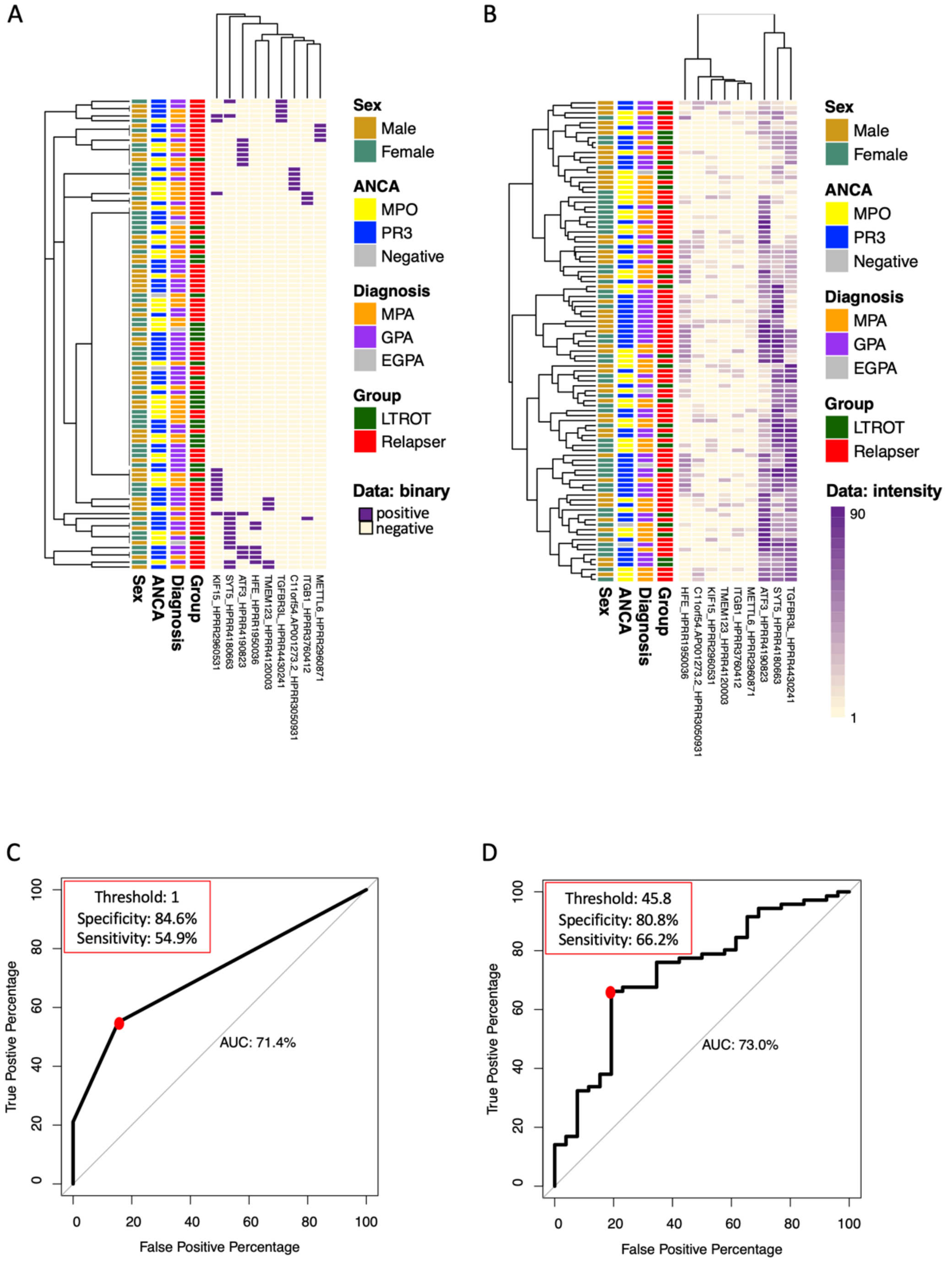


**Supplementary Figure S3. Distribution of the nine selected autoantibodies and classification of LTROTs and frequent relapsers.**
In the upper part of the figure, heatmaps based on binary (A) and normalized intensities [AU] (B) showing the distribution of the nine selected autoantibodies. In the lower part, ROC curves representing the accuracy of the nine antigen panel to separate LTROTs and frequent relapsers based on binary (C) and intensity signals (D).

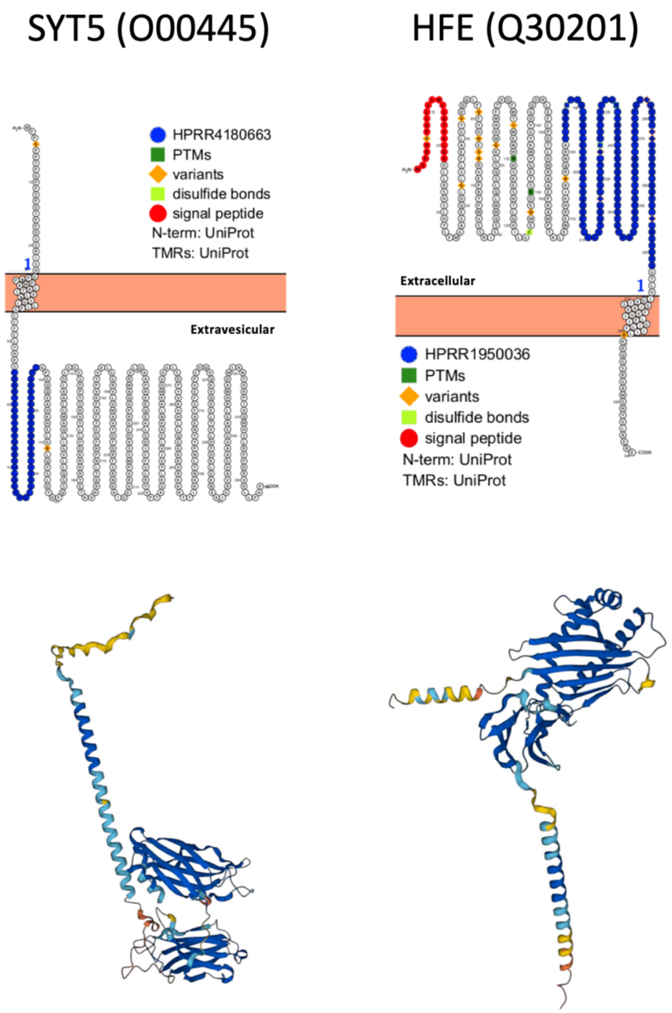


**Supplementary Figure S4. SYT5 and HFE antigen sequence coverage and full-length protein structure.** Top panels: the images were obtained using Protter version 1.0. The part of the protein sequence covered by the protein fragments included in our test is represented in blue for each of the targets. Bottom panels: predicted protein structures from the Alphafold database.
